# Multimorbidity representation via graph learning: A population-based study on hepatosplenic conditions in schistosomiasis-endemic areas of rural Uganda

**DOI:** 10.1101/2024.10.01.24314714

**Authors:** Yin-Cong Zhi, Simon Mpooya, Narcis B. Kabatereine, Betty Nabatte, Christopher K. Opio, Goylette F. Chami

## Abstract

**Background:** The global burden of multimorbidity is increasing yet poorly understood, owing to insufficient methods available for modelling complex systems of conditions. In particular, hepatosplenic multimorbidity has been inadequately investigated.

**Methods:** From 17 January to 16 February 2023, we examined 3186 individuals aged 5-92 years from 52 villages across Uganda within the SchistoTrack Cohort. Point-of-care B-mode ultrasound was used to assess 45 hepatosplenic conditions. Three graph learning methods for representing hepatosplenic multimorbidity were compared including graphical lasso (GL), signed distance correlations (SDC), and co-occurrence. Graph kernels were used to identify thresholds of relevant condition inter-dependencies (edges). Graph neural networks were applied to validate the quality of the graphs by assessing their predictive performance. Clinical utility was assessed through medical expert review.

**Findings:** Multimorbidity was observed in 54·65% (1741/3186) of study participants, who exhibited two or more hepatosplenic conditions. Conditions of mildly fibrosed vessels were most frequently observed (>14% of individuals). Percentage thresholds were found to be 50·16% and 64·46% for GL and SDC, respectively, but could not be inferred for co-occurrence. Thresholded GL and SDC graphs had densities of 0·11 and 0·17, respectively. Both thresholded graphs were similar in predictive utility, although GL produced marginally higher AUCs under certain experiments. Both GL and SDC had significantly higher AUCs than co-occurrence. Numerous conditions were predicted with perfect sensitivity using both GL and SDC with graph convolutional network with five input conditions.

**Interpretation:** The most common method for multimorbidity (co-occurrence) provided an uninformative representation of hepatosplenic conditions with respect to sparsity and predictive performance. More clinically useful graphs were computed when algorithms consisted of statistical assumptions, such as graphical lasso. Future work could apply the pipeline developed here for clinically relevant multimorbidity representations.

**Funding:** NDPH Pump Priming Fund, John Fell Fund, Robertson Foundation, UKRI EPSRC (EP/X021793/1).

## Introduction

The burden of multimorbidity is growing worldwide with an estimated pooled prevalence of at least 33% across high and low-middle income countries.^1^ Multimorbidity is defined as the co-occurrence of two or more chronic health conditions within an individual. Individuals with multimorbidity often are of low socioeconomic status, have greater number of years lived with disability, and experience early mortality.^2^ Hepatosplenic diseases are a particularly complex multimorbidity problem in sub-Saharan Africa with diverse causes ranging from infectious pathogens to non-communicable aetiologies.^3^ There are unique challenges posed by multimorbidity to conventional medical curricula and constrained health systems that cannot be solved by studies focusing on single conditions or diseases. For medical training, there is a need to move from more specialist to generalist medicine, and understand how to provide guidance given the intractability of creating guidelines needed for every possible set of co-occurring conditions. For health systems, there are issues of polypharmacy, misdiagnosis, mistreatment, and more frequent and possibly redundant, costly patient visits. Fundamentally, there remain serious challenges for accurate representation of what multimorbidity exists or will develop in a population.^4^

The epidemiology of multimorbidity has been studied as simplified problems, with well established methods overlooking the inter-dependencies between health conditions to focus on aggregate outcomes. The most common method of classifying individuals as multimorbid is simply counting two or more observed conditions from a predefined, non-exhaustive set of chronic conditions.^2^ This approach ignores how co-occurrence of two disorders arise, which may be by chance, or due to actual shared aetiologies. It is inevitable that the more conditions considered, the more likely an individual will be classified as multimorbid. Meanwhile, factorisation and dimensionality reduction have been considered to model shared underlying patient characteristics of multimorbidity.^5^ However, dimensionality reduction diminishes the discriminative information available from condition inter-dependencies, neglecting the differences between individuals and only allowing clinicians to infer multimorbidity over a homogeneous population.

Graphs can be used to represent the inter-dependencies between conditions and the overall structure of multimorbidity, retaining information that is unique to the patient or diverse populations. The simplest and most common graph construction method is to connect two conditions based on the frequency of co-occurrence.^6–8^ These graphs have a strong assumption that every co-occurrence is equally important and should be considered in the wider multimorbidity graph, inferring relatedness even when conditions manifest together by chance. Graphs also have been learned using pairwise metrics, including t-tests, relative risk, cross entropy loss, cosine similarity, and log odds.^3, 9–13^ However, there currently are no validated decision rules to evaluate the choice of metric or assess its suitability for different multimorbidity problems. Graphical models constitute a different class of graph learning algorithms underpinned by distributional assumptions on the data,^13–15^ wherein relationships between nodes are represented by probabilistic likelihoods. Graphical models can broadly be classified into two types: directed acyclic Bayesian networks and symmetric and undirected Markov networks. Generally, graphical models excel in capturing hierarchical data structures, but can be computationally expensive. As such, edges often are found through search algorithms designed to not exhaustively consider all possibilities, to reduce run time at the expense of potentially sub-optimal graphs. Despite the numerous applications of graphs in clinical studies, there is a notable lack of investigation into the quality of the multimorbidity graphs. Currently graphs are constructed using unvalidated algorithms, often without proper thresholding, and without comparison to alternative algorithms. There is an urgent need to understand hepatosplenic multimorbidity in rural, low-income settings where identification and management strategies are lacking within local health systems. Hepatosplenic multimorbidity in sub-Saharan Africa is complex and often arises due to chronic infections such as hepatitis B/C and parasitic blood flukes of *Schistosoma mansoni,* as well as concurrent alcohol use or aflatoxin exposure.

We assessed 45 hepatosplenic conditions using point-of-care ultrasound to examine 3186 individuals in rural Uganda. We compared algorithms from three families of graph learning with different levels of statistical assumptions to represent complex hepatosplenic multimorbidity. We identified decision rules for thresholding multimorbidity graphs while accounting for the level of morbidity in the population. The quality of the graphs were evaluated based on their utility in multimorbidity prediction, and their ability to uncover insights for medical interpretation. Here we answer the question, how can multimorbidity be assessed and validated in a manner that provides confidence for clinical decision-making?

## Methods

### Participants

This study was conducted within the SchistoTrack prospective cohort^16^ during the first annual follow-up between 17 January and 16 February 2023. 1952 households were randomly sampled from 52 villages across Buliisa, Pakwach, and Mayuge Districts of Uganda; 38 of the villages were sampled in the baseline of 2022.^17^ One child aged 5-17 years and one adult aged 18 years or older were selected by the household head or spouse and invited for clinical assessments. 3224 individuals were clinically assessed. 3186 of 3224 individuals had non-missing ultrasound data and were analysed.

### Hepatosplenic outcomes

We obtained hepatosplenic conditions by point-of-care ultrasound. Philips Lumify C5-2 curved linear array transducers were used with the Philips Lumify Ultrasound Application v3·0 on Lenovo 8505-F tablets with Android 9 Pie. Lossless DICOM images and videos were saved for quality assurance.^17^ A number of indicators were measured including focal and diffuse liver fibrosis patterns, liver surface irregularities, caudal liver edge assessments, fatty and cirrhotic livers, liver and spleen organometry, portal vein dilation or restriction, portosystemic collaterals, ascites, gall bladder obstruction, among others. For the left and right liver lobes, spleen, and portal vein diameter, we measured organometry against an internal healthy reference population standardised by height. We assessed a total of 45 hepatosplenic conditions. Detailed definitions are in the supplementary methods of the Appendix Page 1.

### Population selection

While individuals who were healthy or had only one condition were often excluded from studies on multimorbidity (e.g.^6, 11, 18^), here, all participants were examined, and the similarity or lack thereof between graphs learned across three populations were compared including a mixed population (excluding no one), a morbid population where individuals had at least one condition, and a multimorbid population where individuals had two or more conditions (the conventional population for multimorbidity studies). We refer to a full population henceforth when all participants from the mixed population are used in analyses without splitting the dataset for training and testing.

### Graph learning algorithms

We chose graph learning algorithms from three families of graphs characterized by varying levels of statistical assumptions. Henceforth the conditions are referred to as nodes and the inter-dependencies as edges in the graph. Negative edges were excluded as they arose predominantly due to mutually exclusive conditions or the absence of conditions. As a baseline reference, we used co-occurrence, where edge weights were determined by the total number of individuals that exhibited both conditions concurrently. Hence, edge weights therefore depended on the size of the selected population for analysis. This method lacks any explicit statistical justification and may include edges from only one person and operates under the assumption that each additional co-occurrence contributes equally to the edge weight while ignoring chance.

As an alternative to co-occurrence, we considered the hierarchical correlation of signed distance correlation (SDC) where the coefficients were the edge weights.^19^ This method combined distance correlation and Pearson correlation between two inputs as

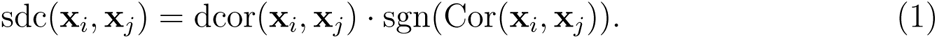

The distance correlation was computed as

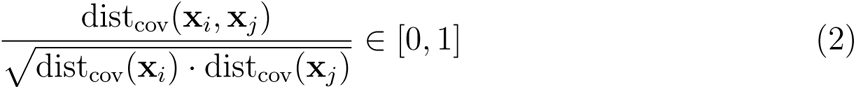

where *L*_2_ norm 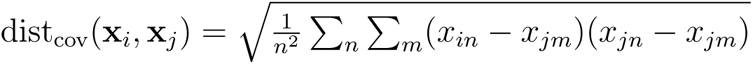, which allowed detection of non-linear dependencies between the data not possible with a simple Pearson correlation. With distance correlation being restricted to positive values, the sign from Pearson correlation was introduced to identify and remove negative correlations as described in.^19^

Moving beyond pairwise metrics to graphical approaches, graphical lasso (GL)^20^ was applied, which assumed the data followed a multi-variate Gaussian distribution and minimized the negative log-likelihood with a sparsity term. Compared to typical graphical models, graphical lasso has advantages in that there exist computationally efficient solutions, and integrated sparsity. The objective function consisted of

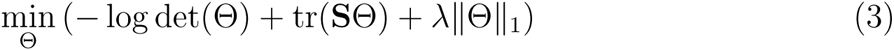

where ϴ was the precision matrix (inverse covariance) used to construct the graph, *-*log det(ϴ) +tr(**S**ϴ) came from the multi-variate Gaussian negative log-likelihood with sample empirical covariance **S**. The *||*ϴ||_1_ term induced sparsity by penalizing the magnitude of the entries in ϴ, while *λ* was the tuning parameter that controlled the trade-off between the log-likelihood and the sparsity term selected through 5-fold cross-validation using the GraphLassoCV package from sklearn 1·5·0 in Python 3·9.

### Thresholding via maximising graph dissimilarity

Thresholding was applied to remove weak connections that may be uninformative and potentially influenced by noise in the data. The algorithms we considered produced different magnitudes of edge weights, therefore percentage thresholding was used to determine the appropriate cutoff. We determined the best threshold by maximising the structural difference between graphs generated on the three populations used to represent multimorbidity. Graph kernels were used to measure structural similarity between two graphs,^21^ where the lower the value the more different the two graphs. We considered three graph kernels from the GraKeL library v0·1·10 in Python 3·9, Weisfeiler-Lehman,^22^ Subgraph matching,^23^ and Neighborhood hash,^24^ details can be found in Appendix Page 5.

We measured the similarity between graphs learned on every combination of the three populations. Kernels were computed over graphs constructed from 500 random samples (to obtain standard deviations) where each sample consisted of a uniform probability random sample of 50% of the study population. The optimal threshold was the location of the minimum kernel value. We averaged over the three kernels and across the three possible population comparisons. Each threshold from each algorithm was applied to the sample graphs to obtain the thresholded graphs, as well as the final graphs produced from each algorithm when re-learned over all participants.

### Algorithm comparison via predictive modelling with GNNs

To evaluate the quality of the graphs from each algorithm, we utilized graph neural networks (GNNs) for the task of multimorbidity prediction. Given the observed status of *m* conditions in an individual, we predicted the status of the full set of 45 conditions assessed in this study. This problem could be viewed as utilizing a partially observed graph representing a scenario of when not all conditions are diagnosed in an individual. We only considered 10 training splits in this experiment, using the first 10 seeds from the previous experiment. 50% of participants were used to train the GNN and the remaining 50% were held out as test set. For each split we randomly selected a subset of *m* conditions to predict both the set of observed conditions and a wider set of unobserved conditions. This approach was taken to represent the problem of where some conditions are known to occur in a population yet there is a need to predict statuses in new patients. The status of each condition was binary, making this a vectorial binary classification over the 45 conditions, and we evaluated performance by AUC, sensitivity, and specificity. All GNNs were set to two layers and a fixed width to allow for comparison across GNNs. Three architectures of graph convolutional network (GCN),^25^ graph attention network (GAT),^26^ and sample and aggregate (GraphSAGE)^27^ were chosen based on their spatial usage of the graph and ease of interpretability. Details can be found in Appendix Page 5. We applied the GNNs to the thresholded graphs with the number of condition inputs *m* = 5, 10*,…,* 25. In this experiment, we also present predictions for only unobserved conditions to represent when new diagnoses need to be evaluated within a patient. As validation analyses, we also varied the threshold to test and compare against the optimal thresholds, and, fixing *m* = 5, applied the GNNs to populations with different levels of morbidity (full population, morbid, and multimorbid), to examine the effect of excluding individuals from the study population.

### Clinical validation

To clinically interpret the graphs and GNNs, the relational information were reviewed by experts including a clinical epidemiologist, and a local sonographer and gastroenterologist to provide insights into the complex root mechanisms of each condition and biological plausibility of inter-dependencies. AUC, sensitivity, and specificity were calculated for each condition averaged over the 10 splits. For sensitivity and specificity, a universal cut-off was selected based on the highest of the two quantities combined over the 45 conditions. One-hop neighbours of the most reliably predicted conditions (highest AUC) were examined to assess their direct influences to compare to standard practice. Global graph properties were examined to assess improvements in sparsity between thresholded and non-thresholded graphs as well as to assess graph stability for clinical decision-making.

## Results

### Observed hepatosplenic conditions

All 45 hepatosplenic conditions were observed at least once in the study population. The most observed conditions were mildly fibrosed vessels, including prominent peripheral rings (462/3186, 14·50%) and prominent pipe stems (456/3186, 14·31%) indicative of early stage periportal fibrosis (Table. 1). Only 18·05% (575/3186) of participants did not exhibit any of the hepatosplenic conditions and 27·31% (870/3186) of individuals exhibited only one condition. Most of the study participants were multimorbid with 54·65% (1741/3186) of individuals with two or more conditions. The median number of conditions across all participants was two (inter-quartile range 1-3). All conditions co-occurred with another condition within at least one person.

### Learned graphs and thresholds

GL and SDC both consistently produced the strongest edge between prominent peripheral rings and prominent pipe stems. These conditions were two different cross-section views of the same pathology and the most observed co-occurrence with a frequency of 437 of 3186 participants. Other regularly observed edges across the samples for each algorithm and population are in Appendix Fig. S1, S2, & S3. Fig. 2 presents the thresholding analysis using graph kernels. The final thresholds were 50·16% for graphical lasso and 64·46% for signed distance correlation.

**Figure 1:**
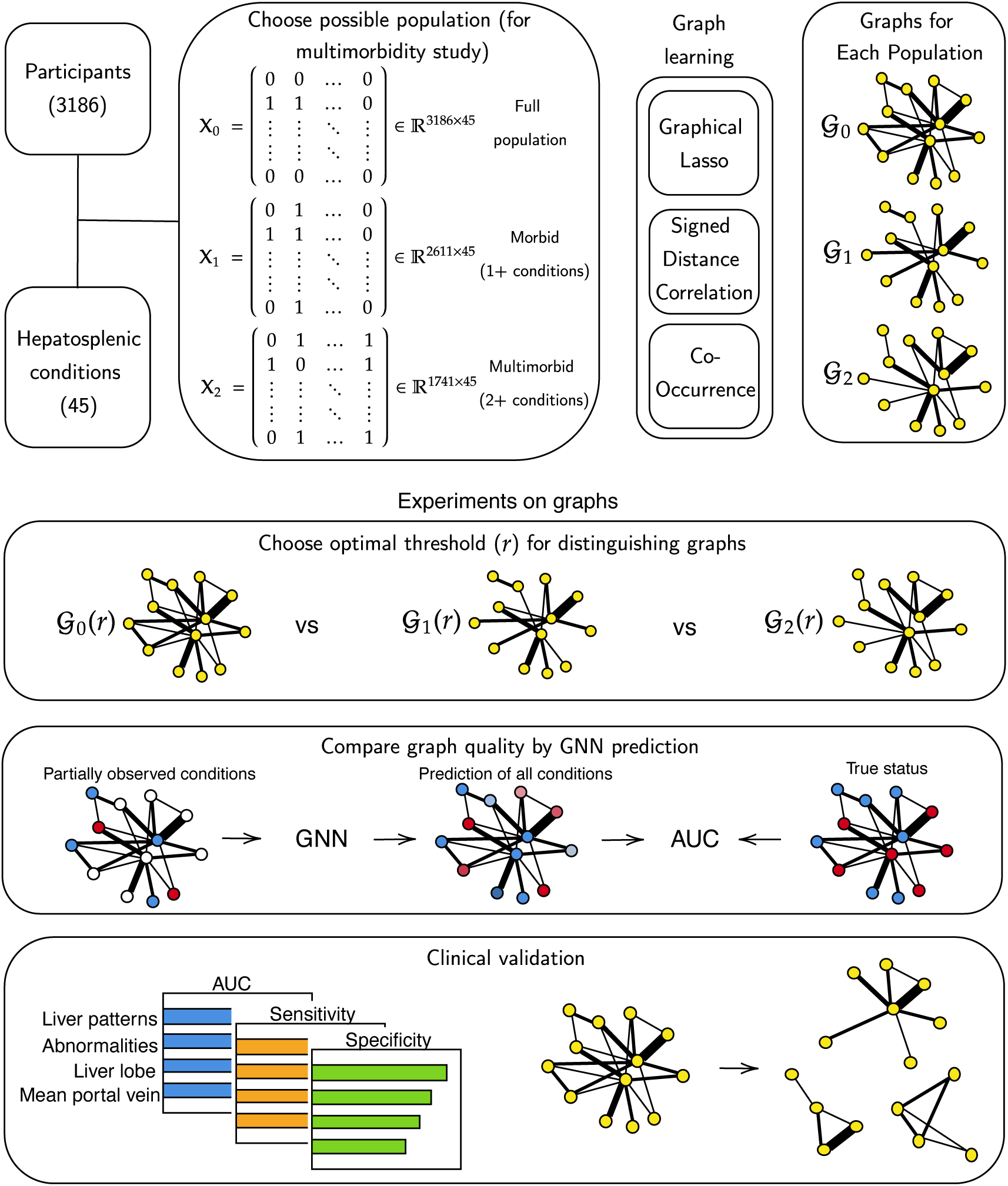
Overview of the graph learning pipeline.

**Figure 2:**
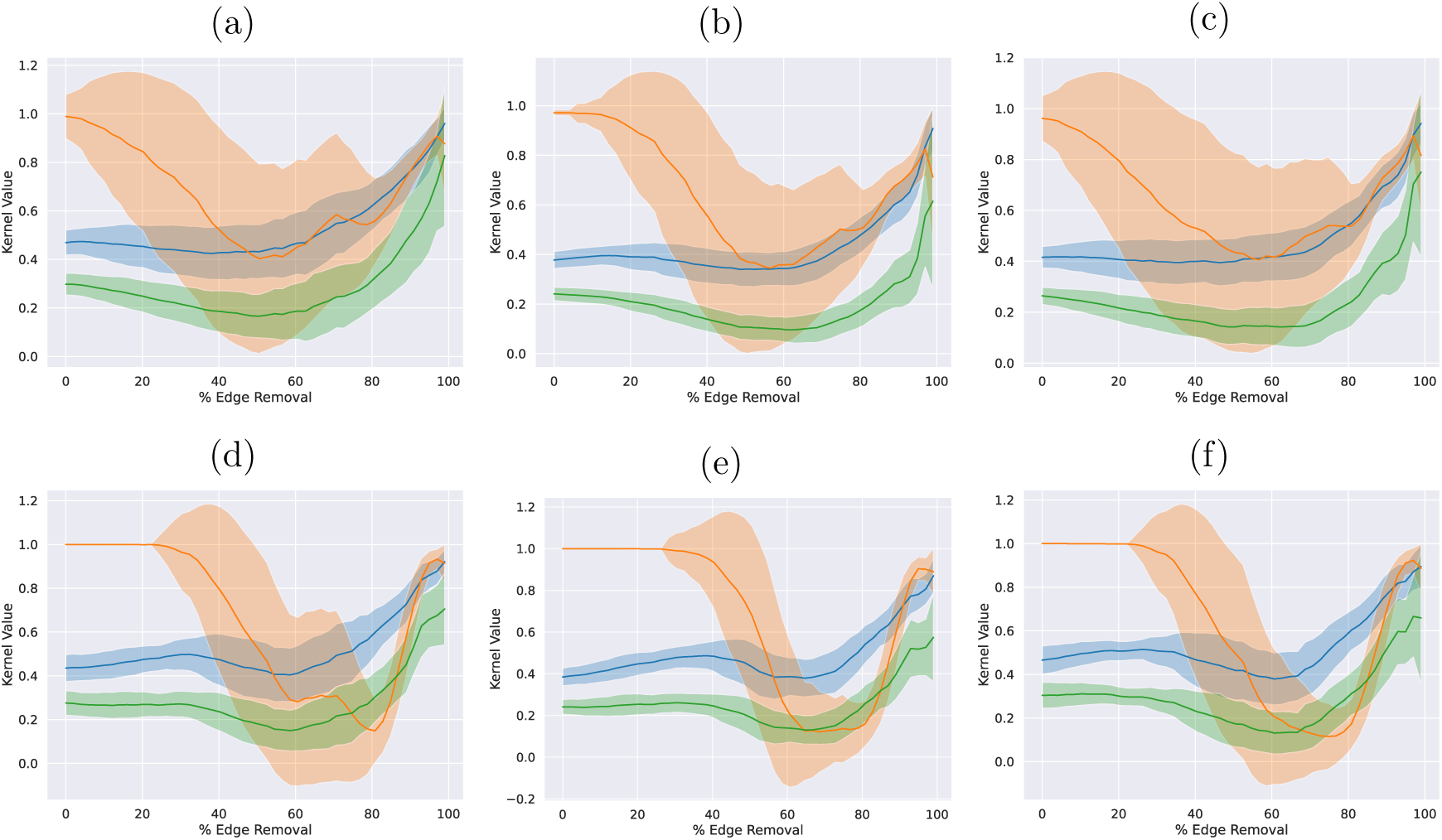
Graph kernels results. Blue: Weisfeiler Lehman, Orange: Subgraph Matching, Green: Neighborhood Hash. (a) Graphical lasso full vs morbid. (b) Graphical lasso full vs multimorbid. (c) Graphical lasso morbid vs multimorbid. (d) Signed distance correlation full vs morbid. (e) Signed distance correlation full vs multimorbid. (f) Signed distance correlation morbid vs multimorbid. Plots show the similarity measures between graphs learned the full population, morbid (1+ condition), and multi-morbid people (2+ conditions) using graph kernels. This experiment cannot be applied to co-occurrence graphs as they are identical when constructed from any of the three populations.

Global properties of the 500 thresholded graphs are presented in Table 2 (non-thresholded graphs in Appendix Table S1). Despite a higher percentage threshold, SDC was denser than GL, and had a more discernible degree structure fitting to a log-normal distribution while GL did not fit common degree distributions. SDC and GL had edge densities of 0·17 and 0·11 with average degree of 7·29 and 4·84, respectively. There was a 54% overlap in edges between the two graphs when considering the union of edges. GL maintained similar global properties between the final graph and the training samples, whereas SDC was sensitive to the sample size, and the final and training samples from SDC were significantly different in a number of statistics. Concerning co-occurrence, there were no identifiable thresholds as co-occurrence edges were only defined by multimorbid participants and excluding the healthy and morbid did not change the graph. Consequently, co-occurrence had the highest edge density of 0·55 and average degree of 24·23 (exhibiting a power law degree distribution) where all nodes were connected in a single component (Appendix Table S1). The final thresholded graphs computed over all participants are shown in Fig. 3, whereas the unthresholded co-occurrence graph is presented in Appendix Fig. S6 along with the unthresholded GL (Appendix Fig. S4) and SDC (Appendix Fig. S5).

**Figure 3:**
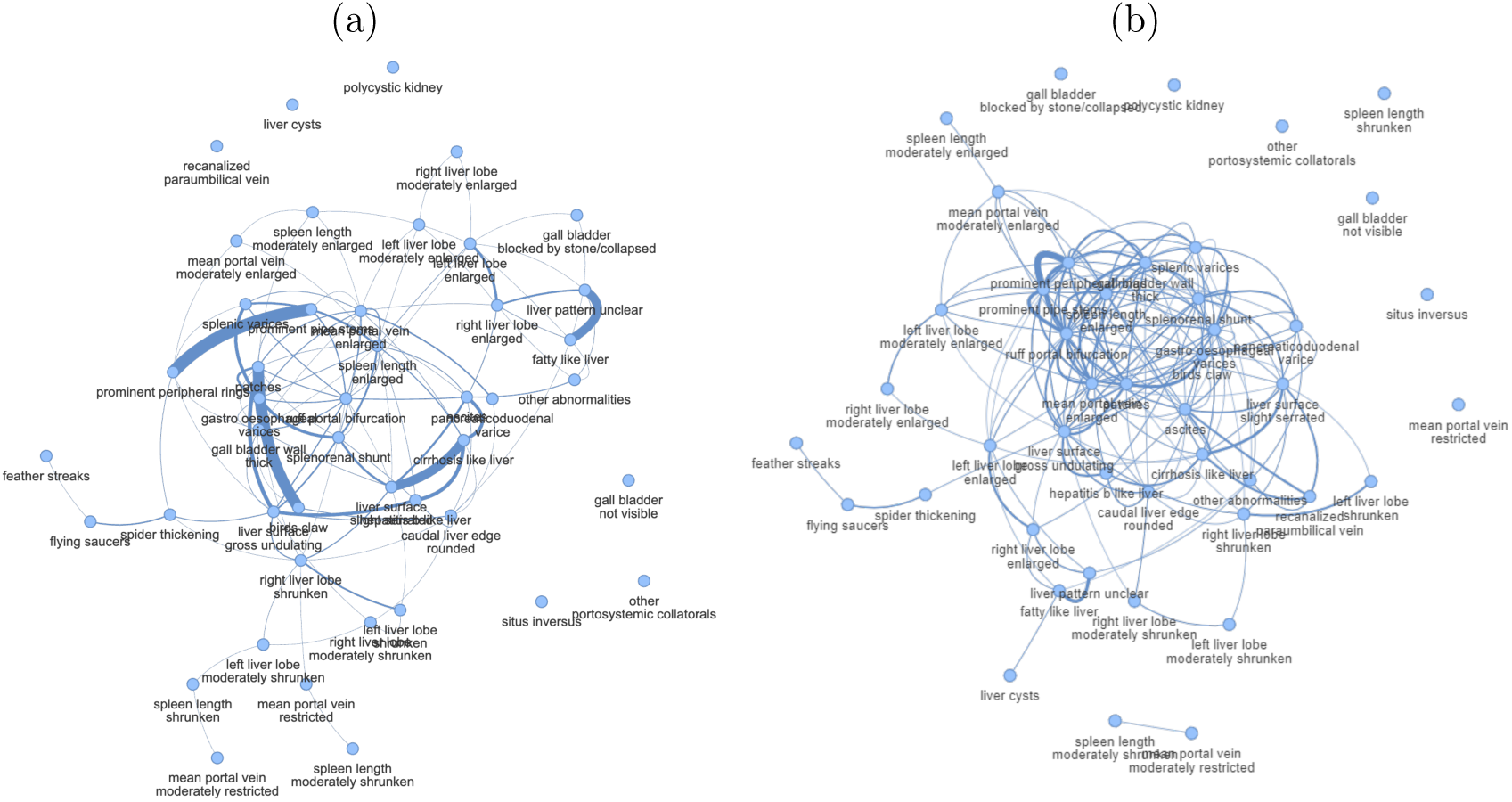
Final graphs using optimal percentage thresholds. Each graph is learned on the full dataset using the average thresholds found from the graph kernels experiment for (a) graphical lasso (50·16%) (b) signed distance correlation (64·46%).

**Table 1:** List of conditions and the number of participants. *Outcomes in italic* indicate the healthy form and were not included in the multimorbidity graph.

| Variable | Outcomes | # All Participants (%)<br>Total = 3186 | # 1+ Conds (%)<br>Total = 2611 | # 2+ Conds (%)<br>Total = 1741 |
| --- | --- | --- | --- | --- |
| Liver Patterns | *Normal* | 2340 (73.45) | 1765 (67.60) | 949 (54.51) |
|  | Unclear | 15 (0.47) | 15 (0.57) | 15 (0.86) |
|  | Feather Streaks | 239 (7.50) | 239 (9.15) | 213 (12.23) |
|  | Flying Saucers, Starry Sky | 195 (6.12) | 195 (7.47) | 179 (10.28) |
|  | Spider Thickening | 53 (1.66) | 53 (2.03) | 48 (2.76) |
|  | Prominent Peripheral Rings | 462 (14.50) | 462 (17.69) | 459 (26.36) |
|  | Prominent Pipe Stems | 456 (14.31) | 456 (17.46) | 455 (26.13) |
|  | Ruff, Portal Bifurcation | 185 (5.81) | 185 (7.09) | 182 (10.45) |
|  | Patches (occluded, bright white vessels) | 45 (1.41) | 45 (1.72) | 45 (2.58) |
|  | Bird's Claw | 8 (0.25) | 8 (0.31) | 8 (0.46) |
| Other Abnormalities | *None* | 2951 (92.62) | 2376 (91.00) | 1525 (87.59) |
|  | Cirrhosis-like Liver | 12 (0.38) | 12 (0.46) | 12 (0.69) |
|  | Fatty-like Liver | 65 (2.04) | 65 (2.49) | 61 (3.50) |
|  | Heptatitis-B-like Liver | 65 (2.04) | 65 (2.49) | 62 (3.56) |
|  | Polycystic Kidney Disease | 2 (0.06) | 2 (0.08) | 2 (0.11) |
|  | Liver Cysts | 4 (0.13) | 4 (0.15) | 3 (0.17) |
|  | Situs Inversus | 1 (0.03) | 1 (0.04) | 1 (0.06) |
|  | Other | 47 (1.48) | 47 (1.80) | 44 (2.53) |
| Liver Surface | *None* | 3135 (98.40) | 2560 (98.05) | 1691 (97.13) |
|  | Slight/Serrated | 17 (0.53) | 17 (0.65) | 16 (0.92) |
|  | Gross/Undulating | 34 (1.07) | 34 (1.30) | 34 (1.95) |
| Caudal Liver Edge | *Sharp* | 2908 (91.27) | 2333 (89.35) | 1495 (85.87) |
|  | Rounded | 278 (8.73) | 278 (10.65) | 246 (14.13) |
| Left Liver Lobe | *Normal* | 2212 (69.42) | 1637 (62.70) | 923 (53.01) |
|  | Moderately Enlarged | 428 (13.43) | 428 (16.39) | 364 (20.91) |
|  | Moderately Shrunken | 357 (11.21) | 357 (13.67) | 285 (16.37) |
|  | Enlarged | 85 (2.67) | 85 (3.26) | 81 (4.65) |
|  | Shrunken | 104 (3.26) | 104 (3.98) | 88 (5.05) |
| Right Liver Lobe | *Normal* | 2198 (68.99) | 1623 (62.16) | 929 (53.36) |
|  | Moderately Enlarged | 410 (12.87) | 410 (15.70) | 326 (18.72) |
|  | Moderately Shrunken | 412 (12.93) | 412 (15.78) | 330 (18.95) |
|  | Enlarged | 52 (1.63) | 52 (1.99) | 51 (2.93) |
|  | Shrunken | 114 (3.58) | 114 (4.37) | 105 (6.03) |
| Mean Portal Vein | *Normal* | 2156 (67.67) | 1581 (60.55) | 902 (51.81) |
|  | Moderately Enlarged | 425 (13.34) | 425 (16.28) | 360 (20.68) |
|  | Moderately Restricted | 388 (12.18) | 388 (14.86) | 284 (16.31) |
|  | Enlarged | 168 (5.27) | 168 (6.43) | 154 (8.85) |
|  | Restricted | 49 (1.54) | 49 (1.88) | 41 (2.35) |
| Portosystemic Collaterals | *Not Detected* | 3152 (98.93) | 2577 (98.70) | 1707 (98.05) |
|  | Splenic Varices | 12 (0.38) | 12 (0.46) | 12 (0.69) |
|  | Gastro-oesophageal Varices | 11 (0.35) | 11 (0.42) | 11 (0.63) |
|  | Pancreaticoduodenal Varices | 5 (0.16) | 5 (0.19) | 5 (0.29) |
| | Entirely Recanalized Paraumbilical Vein $\geq 3\text{mm}$ | 1 (0.03) | 1 (0.04) | 1 (0.06) |
|  | Splenorenal Shunt | 13 (0.41) | 13 (0.50) | 13 (0.75) |
|  | Other | 1 (0.03) | 1 (0.04) | 1 (0.06) |
| Ascites | *Not Detected* | 3174 (99.62) | 2599 (99.54) | 1730 (99.37) |
|  | Yes | 12 (0.38) | 12 (0.46) | 11 (0.63) |
| Gall Bladder Visible | No | 30 (0.94) | 30 (1.15) | 19 (1.09) |
|  | Yes, but blocked by stone or collapsed | 117 (3.67) | 117 (4.48) | 101 (5.80) |
|  | *Yes, Clearly Visible* | 3090 (95.39) | 2464 (94.37) | 1621 (93.11) |
| Gall Bladder Wall | *Normal* | 2939 (92.25) | 2364 (90.54) | 1512 (86.85) |
|  | Thick | 247 (7.75) | 247 (9.46) | 229 (13.15) |
| Spleen Length | *Normal* | 2188 (68.68) | 1613 (61.78) | 938 (53.88) |
|  | Moderately Enlarged | 387 (12.15) | 387 (14.82) | 315 (18.09) |
|  | Moderately Shrunken | 392 (12.30) | 392 (15.01) | 289 (16.60) |
|  | Enlarged | 182 (5.71) | 182 (6.97) | 168 (9.65) |
|  | Shrunken | 37 (1.16) | 37 (1.42) | 31 (1.78) |

**Table 2:** Sample and final graph statistics. Sample graph statistics were computed over 500 samples at optimal thresholds, and final graphs were learned on training and testing sets combined. Final graphical lasso and signed distance correlation graphs share 92 edges, and 34 nodes in their largest components. Degree distribution of none indicated that the graphs did not fit any of exponential, power law, or log-normal.

|  | Graphical Lasso Samples | Final GL Graph | Signed Distance Correlation Samples | Final SDC Graph |
| --- | --- | --- | --- | --- |
| Number of Edges | 110.99 ± 4.60 | 109 | 148.37 ± 4.84 | 164 |
| Average Degree | 4.93 ± 0.20 | 4.84 | 6.59 ± 0.22 | 7.29 |
| Average Clustering Coefficient | 0.21 ± 0.03 | 0.26 | 0.37 ± 0.03 | 0.39 |
| Diameter | 6.30 ± 0.93 | 6 | 6.23 ± 0.99 | 7 |
| Edge Density | 0.11 ± 0.00 | 0.11 | 0.15 ± 0.00 | 0.17 |
| Largest Component Size | 39.64 ± 1.65 | 39 | 36.37 ± 1.81 | 36 |
| Number of Isolated Nodes | 4.73 ± 1.27 | 6 | 6.83 ± 1.63 | 7 |
| Assortativity | 0.12 ± 0.08 | 0.12 | 0.25 ± 0.07 | 0.32 |
| Largest Component Spectral Gap | 0.66 ± 0.30 | 0.32 | 0.04 ± 0.04 | 0.03 |
| Degree Distribution | None | None | Log-normal | Log-normal |

### Graph quality evaluation by GNN predictive analysis

Fig. 4 compares the thresholded graphs and GNNs used for multimorbidity prediction (further validation by varying thresholds shown in Appendix Fig. S8 – S18). The prediction problem, which used a partially observed graph, represented a scenario for when only some conditions are observed in a patient and there is a need to also predict undiagnosed conditions. For example, when only 11% (5/45) of conditions were observed by the GCN, the average AUC was 0·75, 0·73, and 0·56 for GL, SDC, and co-occurrence, respectively. Out of the 10 training splits, the highest test AUC (0·78) was achieved when indicators of liver fibrosis, fatty livers, portal vein enlargement possibility indicative of portal hypertension, and hypersplenism were observed. The worst performing split included rounded liver edges, recanalized paraumbilical veins, gall bladder injuries, and abnormal spleen organometry (AUC of 0·69). GL and SDC performed similarly if not better when AUCs were evaluated over only unobserved conditions. Generally, GL and SDC produced similar AUCs with all three GNNs. Although marginally, GL performed best when considering consistent differences to SDC across varying numbers of condition inputs for GAT and GraphSAGE. Co-occurrence on the other hand, with 0% thresholding, was the worst performing graph when evaluated with GCN and GAT, although co-occurrence showed comparable performance for GraphSAGE. Similar results on unobserved conditions only are presented in Appendix Fig. S7. All three GNNs performed similarly for GL and SDC when tested over each of the mixed, morbid, and multimorbid populations; whereas the co-occurrence graph construction was only possible for a multimorbid population (see Appendix Fig. S9-S18)

**Figure 4:**
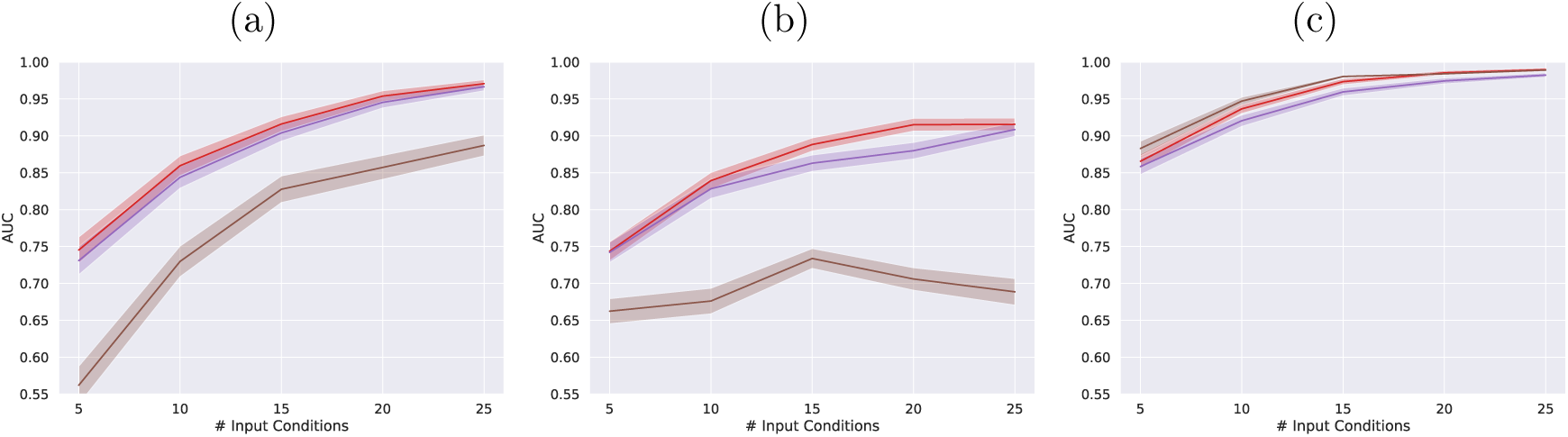
Multimorbidity prediction with varying number of inputs. (a) GCN, (b) GAT, & (c) GraphSAGE, the model uses graphs with optimal thresholds, Red: graphical lasso 50·16%, Purple: signed distance correlation 64·46%, Brown: Cooccurrence 0%. An optimal threshold could not be found for the co-occurrence graph, so 0 threshold was used instead; this matched with what had been done in the literature.^6, 7^ All AUCs were averaged over 10 training-testing splits.

### Hepatosplenic multimorbidity

Fig. 5 presents the GCN model performance broken down for each condition using GL due to the marginally superior performance when compared to SDC. For conditions not exhibited by anyone in the test set, AUC cannot be computed, so we observed the training set for an indication of their performances (Appendix Fig. S19, S20, & S21). With a range of AUCs from 0·992-0·975, the conditions best predicted included moderate periportal fibrosis (prominent pipe stems) and severe conditions of portal fibrosis extending to the liver capsule (bird’s claw pattern), ascites, and splenorenal shunts. These generally had degrees close to the mean degree of the graph, while the frequency of these conditions vastly varied. Sensitivity and specificity for each condition are shown in Appendix Fig. S22 and S25 (more comprehensive plots presented in Appendix Fig. S22 – S27). Using GL, the GCN model showed higher sensitivity than specificity for 37 of 42 conditions (88%; where three conditions could not be evaluated) with an average sensitivity of 0·91 and average specificity of 0·67. The final graphs from GL and SDC are presented in Figs. 3a and 3b. Biologically relevant information for the hepatosplenic system where, for example, splenic enlargement was one of the most central conditions was produced from both graphs. One-hop neighbourhood subgraphs for the best predicted conditions from the GL are shown in Fig. 6. For example, conditions considered multimorbid with the liver condition of prominent pipe stems, which was predicted with high accuracy (AUC 0·992), included ascites, patches, enlarged mean portal vein, and ruff portal bifurcation. Conditions of situs inversus, some portosystemic collaterals, polycystic kidneys, and gall bladder not visible appeared in both graphs as isolates, indicating that they were independent to the rest of the conditions.

**Figure 5:**
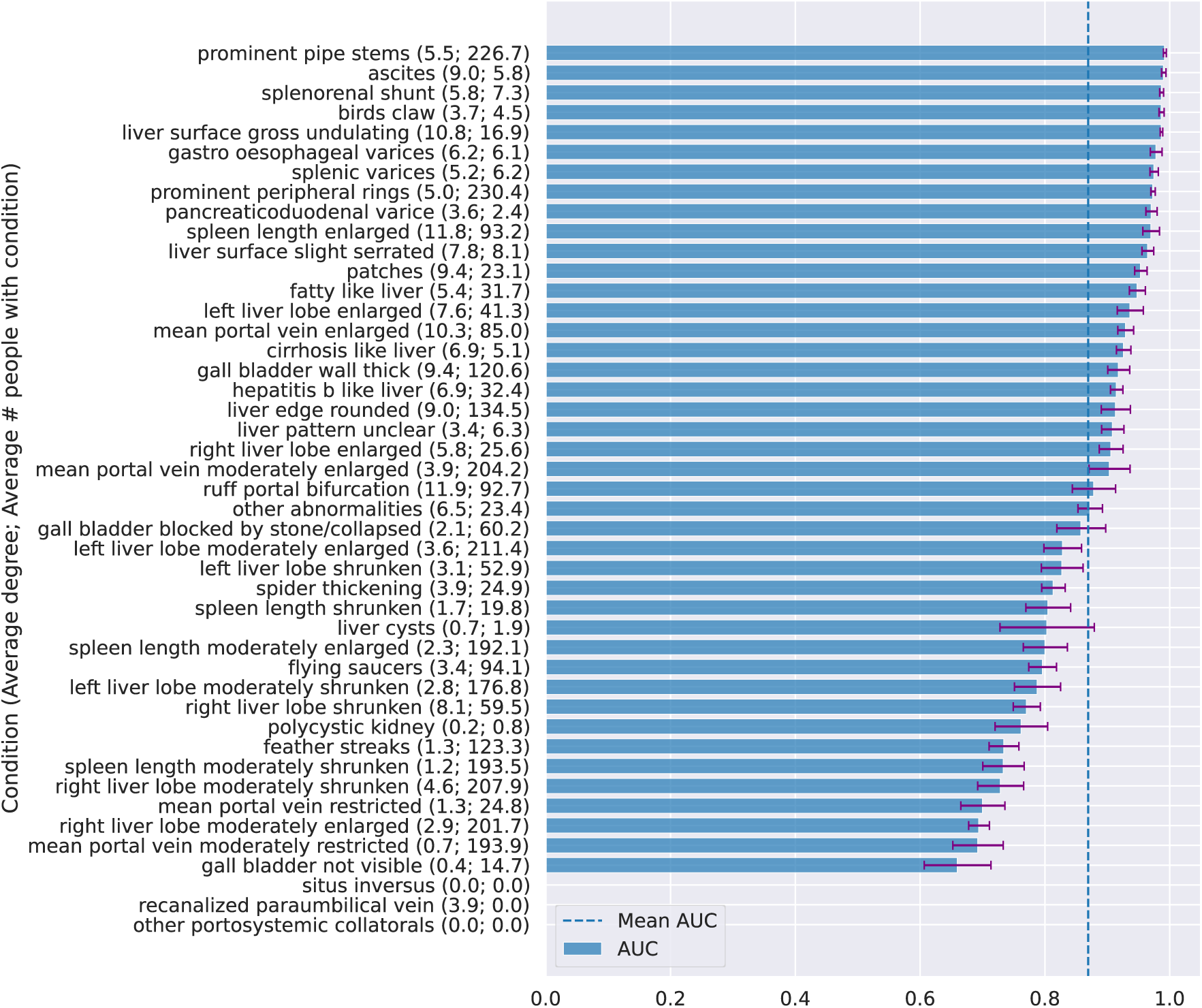
Performance on each condition ordered by AUC. Results were produced by a GCN using graphical lasso at optimal threshold on the test sets. Results on signed distance correlation and co-occurrence can be found in Appendix Fig. S19, S20, & S21. For conditions only observed once, the positive patients were required to be in the training set to run the graph learning algorithms, so the testing set consisted of only one class and AUCs did not exist. For these conditions one can observe the performances on the training sets from the additional results in Appendix Fig. S19, S20, & S21 for an informed indication of performance.

**Figure 6:**
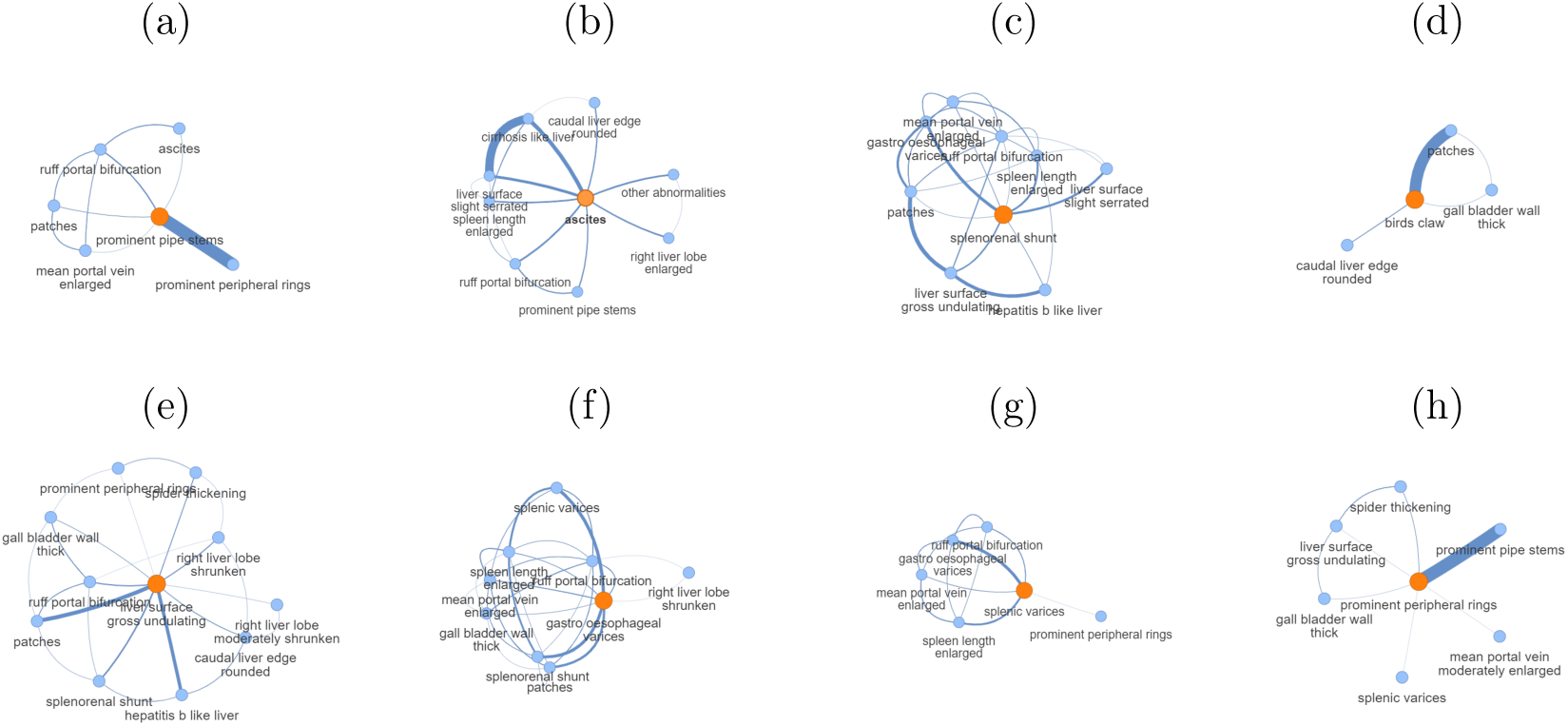
Neighbourhood subgraphs of the conditions with the highest AUCs. Examples are taken from Fig. 5. The subgraphs are taken from thresholded final graphical lasso graph. (a) AUC = 0.992 *±* 0.007. (b) AUC = 0.990 *±* 0.011. (c) AUC = 0.987 *±* 0.010. (d) AUC = 0.987 *±* 0.013. (e) AUC = 0.987 *±* 0.007. (f) AUC = 0.978 *±* 0.030. (g) AUC = 0.975 *±* 0.021. (h) AUC = 0.973 *±* 0.011.

## Discussion

Graph learning is an essential step towards understanding complex multimorbidity. 3186 individuals within the SchistoTrack study in rural Uganda were diagnosed with 45 hepatosplenic conditions using point-of-care ultrasound. Analysing these conditions, we presented a machine learning pipeline to learn clinically useful graphs. We established decision rules for thresholding statistically relevant condition inter-dependencies and evaluated the graphs for multimorbidity prediction. We showed that co-occurrence graphs were poorly suited for problems of multimorbidity. Our study produced sparser, more interpretable graphs using GL and SDC that offered clearer clinical insights for understanding hepatosplenic morbidity in low-income countries.

While co-occurrence has been regularly used to construct multimorbidity graphs,^6–8^ we found co-occurrence graphs to be over-specified (overestimating multimorbidity), dense with little discriminatory information for conditions, and with low predictive utility. Sparser graphs than those observable with co-occurrence enable superior predictive modelling and are easier to interpret.^28^ We showed that thresholding graphs based on maximising structural differences between full, morbid, and multimorbid populations removed weak inter-dependencies (edges) that represented insignificant relations or noise in the data, producing clinically informative sparse multimorbidity graphs. In particular, GL had the lowest edge density and average degree when compared to co-occurrence or SDC. Both GL and SDC detected nodes as isolates, indicating the lack of significant clinical associations with other conditions, while co-occurrence forced inter-dependencies between these conditions that co-existed in a small number of people (often just 1), which is insufficient evidence to identify any aspect of multimorbidity as a public health problem across a population.

The limited utility of co-occurrence was further evident when applied to the task of multimorbidity prediction. Graphs from GL and SDC both improved neural network models by similar margins, and significantly better than co-occurrence in two of the three GNNs. A near complete removal of all graph edges led to the lowest AUCs from each GNN, highlighting the utility of the graphs. These results indicate that multimorbidity could be represented accurately by multiple graphs from inherently different algorithms that had at least some level of statistical assumption. Although co-occurrence performed well with GraphSAGE, GraphSAGE has a sampling framework that is well-suited to denser graphs, as only a subset of neighbours were used from each node. Although GraphSAGE could potentially alleviate the need for thresholding, this model is less interpretable clinically as the stochasticity makes it difficult to identify inter-dependencies for a diagnosed condition, whereas conditions might not be included in the aggregation steps of GraphSAGE despite belonging to the neighbourhood of the condition of interest. Thus, despite the better performance of GraphSAGE,^18^ when building models for predictions with good interpretability, GCN and GAT were better supported here for multimorbidity problems.

Methods of analysing graphs in order to define multimorbidity vary widely. Community detection often is performed on graphs, with each cluster of conditions labelled as multimorbid.^5, 6^ Edge prediction has been explored and multimorbidity has been conceptualized as pairwise relationships between two conditions of interest.^29^ Here we proposed a different approach to understanding multimorbidity that takes into account the entire complex system of hepatosplenic conditions. We characterized multimorbidity by the diagnosis and prediction of multiple conditions, extending far beyond the prediction of two conditions and retaining information specific to each individual condition. Connectivity between two conditions in the learned graph implied the pair were multimorbid. The practical usage of these graphs may be as follows. If a patient was diagnosed with one condition, the sub-graph centred on that condition could be inspected to identify what the patient also is likely to have or what later conditions they are likely to develop. For example, our study suggests that individuals with mild periportal fibrosis might later develop severe conditions indicative of portal hypertension such as ascites, extensive portal fibrosis, and enlarged main portal vein diameters. In the case of multiple positive diagnoses, a clinician may consider using the union of neighbourhoods in our graphs to derive the full set of multimorbidity in a patient.

The GCN had the most interpretable architecture and was therefore analyzed to reveal the best and worst predicted conditions. Conditions with very few degrees or isolate conditions generally had the lowest AUCs. Hence, connectivity was important for modelling multimorbidity and the use of neighborhood information supported more accurate predictions of undiagnosed conditions. Interestingly, the best predicted conditions were not the most connected (hubs) nor necessarily the most frequent (highest population prevalence). When focusing on the prediction of individual conditions, our interpretable GCN model was much better at predicting positive diagnoses rather than the absence of a diagnosis or condition (higher sensitivity than specificity). This trend is to be expected as in clinical practice, with limited time and resources, it often is an insurmountable problem to confirm true negatives without extensive, exhaustive alternative diagnostics such as alternative imaging modalities or biopsies that would have been needed here. Thus, if using our model for hepatosplenic conditions, one may envision confidence in providing treatments when a patient is predicted to have a set of conditions, but we would recommend further clinical review and follow-up if a patient is predicted as unlikely to have a set of conditions. All of which is in line with standard general medicine practice. Moreover, we discovered that no population selection was needed for the study of multimorbidity. GL alleviated the need to focus on only individuals with two or more conditions, which is needed for co-occurrence graphs. Graph learning methods based on statistical assumptions make better use of the full information available from a population, which might enable better prediction for individuals who are yet to develop multimorbidity.

Learning how to accurately represent multimorbidity using graph learning for clinical-decision making opens avenues for more advanced modelling of multimorbidity that incorporates individual patient characteristics into multi-output models (e.g.^30^) where we move beyond current medical practice focusing on one disease per patient. Our work not only revealed the complex system of inter-dependencies for hepatosplenic conditions, but also provides a validated machine learning pipeline for the wider clinical and research community that could be followed to understand multimorbidity as a public health problem across populations.

## Author contributions

Conceptualization: GFC and YCZ. Data curation and validation: YCZ. Formal analysis: YCZ. Investigation, methodology, visualization: YCZ. Writing – original draft: YCZ and GFC. Validation: YCZ, GFC, CKO, and SM. Writing – review and editing: YCZ, GFC, CKO, SM, NBK. Funding acquisition and supervision: GFC. Resources: GFC. Data collection: GFC, SM, NBK.

## Declaration of interest

The authors declare no conflict of interest.

## Data availability

Participant data are not publicly available due to data protection and ethics restrictions related to the ongoing nature of the SchistoTrack Cohort and the easily identifiable sensitive characteristics of the participants.

## Ethics approvals

Data collection and use were reviewed and approved by Oxford Tropical Research Ethics Committee (OxTREC 509-21), Vector Control Division Research Ethics Committee of the Uganda Ministry of Health (VCDREC146), and Uganda National Council of Science and Technology (UNCST HS 1664ES).

## Funding

This work was supported by the NDPH Pump Priming Fund, John Fell Fund, Robertson Foundation Fellowship, and UKRI EPSRC (EP/X021793/1) to G.F. Chami. For the purpose of Open Access, the author has applied a CC BY public copyright licence to any Author Accepted Manuscript version arising from this submission.

## Acknowledgements

We are thankful for the involvement from our study participants, and the SchistoTrack teams especially the surveyors, nurses, sonographers, and laboratory technicians. We also like to thank the Uganda Ministry of Health, local district leaders, focal health workers, and village health teams. Special thanks also to the Oxford team for the fieldwork, data wrangling, everyday discussions, and feedback.

## Research in Context

Graphs are commonly used across the biomedical sciences to study complex relationships from cellular systems to clinical outcomes. There exists a vast array of methods to analyze graphs and real-world networks to understand human health. Yet, the usefulness of any graph depends on the quality of the data and ground truth structure, and most importantly the method of learning the graph structure and validation when no ground truth or references exist. Methods for learning and validating graphs that best represent human multimorbidity have been poorly explored. Multimorbidity is the existence of two or more chronic conditions within an individual, and is rising worldwide especially in low-income countries where there is a double burden of infectious and non-communicable diseases. An urgent example of poorly understood multimorbidity is the complex system of inter-dependencies of hepatosplenic conditions. To identify a graph suitable for clinical-decision making, open questions remain of how to determine the relevance of condition relationships, of how best to choose a threshold for determining what is a significant clinical relationship of public health or individual concern (i.e. deciding the presence of an edge between conditions in the graph), and of how to validate that threshold for understanding or predicting multimorbidity in the absence of any ultimate ground truth given the context of complex, dynamic human diseases. Thus, a pipeline urgently is needed for representing multimorbidity through graph learning. Henceforth, we focus on the complex problem of hepatosplenic multimorbidity as a known complex system with diverse biological causes.

We conducted a systematic search of titles and abstracts in PubMed from database inception to 12 April 2024 using the following search string: (“network*” OR “graph*”) AND (“model*” OR “threshold*” OR “predict*”) AND (“comorbidit*” OR “co-morbidit*” OR “multimorbidit*” OR “multi-morbidit*” OR “multiple conditions” OR “multiple diseases”) AND (“spleen” OR “liver” OR “hepat*”). Our search string yielded 115 studies, from which we removed studies, based on the title and abstract, that did not involved the use of graphs or networks to model and understand multimorbidity data on or relating to the liver, spleen, or both, leaving us with only 25 studies. Given the low number of studies on hepatosplenic diseases, we broadened our search string to (“network*” OR “graph*”) AND (“model*” OR “threshold*” OR “predict*”) AND (“comorbidit*” OR “co-morbidit*” OR “multimorbidit*” OR “multi-morbidit*” OR “multiple conditions” OR “multiple diseases”), but reducing to titles searches only. This search returned 26 publications, of which only one was also in the 25 initially screened hepatosplenic-related studies. We manually reviewed the abstracts and the full texts of these studies and removed nine studies that focused on neural networks only as opposed to studying networks as structures representative of real-world systems. The remaining 17 studies investigated datasets such as patient clinical data, proteins, and genes datasets.

From the combined list of 25 hepatosplenic-related studies and 17 multimorbidity graph modelling (including one study in both categories), no study developed methods to determine graph thresholds or even considered the influence of thresholding on graph quality for the application at hand. From the relevant studies identified, no studies directly reported using data from sub-Saharan Africa. Only one publication compared different graph learning algorithms, but only considered three graphical models, thus comparing from the same family of graphs with the same level of statistical assumptions on graph generation. No studies investigated how to threshold or remove edges from the graph in order to remove inter-dependencies that are likely due to chance and less informative for clinical decision-making. No studies provided means of validating the graph used for biomedical analysis. Published studies often were focused on graphs where there was a naturally occurring ground truth (graph structure) that could be validated with experimental laboratory work (e.g. protein-protein, genes, drug interactions); otherwise, simple co-occurrence was examined where any two conditions reported or observed together for individuals were considered multimorbid without any statistical adjustment of chance co-occurrence. The clinical interpretation of the graphs for multimorbidity were limited to focusing on the directly connected conditions of interest (neighbours of nodes), notably throwing away information from the wider set of conditions within the graph that were not directly connected (or co-reported) instead of incorporating information from the full graph of conditions. No study investigated the use of different populations, i.e. splitting the people studied based on how many conditions observed in each person or questioning whether to include healthy participants in analyses. The focus of studies was on individuals who were already multimorbid, which might limit predicting new multimorbidity development within an individual or population or limit insights into how the wider hepatosplenic system functions. How well the graph aided multimorbidity prediction tasks was not explored in any of the studies identified.

## Added value of this study

The value of our study lies in the insight provided into the complex system of hepatosplenic multimorbidity. Importantly, we provide methods for how to select thresh-olds for multimorbidity graphs more broadly based on the morbidity characteristics of the population and how to assess the quality of the graphs through predictive tasks to aid clinical decision-making. We evaluated a real-world network of hepatosplenic conditions affecting individuals in low-income settings where both infectious and noncommunicable causes are common. Hepatosplenic conditions are known biologically to exhibit strong inter-dependencies and here we extend this knowledge to an identifiable graph structure between liver and spleen conditions commonly observed in rural Uganda. Forty-five hepatosplenic conditions across 3186 individuals aged 5 years and older were investigated within the SchistoTrack cohort that is based in 52 rural communities in Uganda.

We demonstrated the usefulness of considering sparsity and thresholding to improve the quality of the final multimorbidity graph. We presented a set of methods to select relevant edges (associations) between conditions rather than simply retaining all possible edges as commonly practised in existing studies of co-occurrence. Although thresholding has been applied with correlation-based graphs, the exact cut-off in past studies has been generally chosen arbitrarily, and correlation as edge weights often lead to dense graphs. We analyzed and compared three graph learning methods with vastly different statistical assumptions on the data: co-occurrence (no assumption), signed distance correlation (pairwise statistics), and graphical lasso (distributional assumption). Thresholds were inferred based on structural differences, measured by graph kernels, when applied to different population splits consisting of varying levels of morbidity. These were found to be 50·16% for graphical lasso and 64·46% for signed distance correlation. The quality of the graphs were then assessed through multimorbidity predictions evaluated for every condition from graph neural networks (GNNs) where graph convolutional networks (GCN), graph attention networks (GAT), and sample and aggregate (GraphSAGE) were considered that used the thresholded graphs. Thresholded graphical lasso and signed distance correlation graphs performed similarly. For example, graphical lasso, achieved average AUCs from 0·75 – 0·85 across the GNNs when only five of the 45 conditions were observed.

Graphical lasso, which has been rarely used in any multimorbidity graph learning papers, showed properties of stability and sparsity that are ideal for understanding multimorbidity, producing more interpretable bespoke graphs to the population of study, and larger performance improvements when integrated with predictive models. In a study of multimorbidity, often decisions need to be made regarding whether to analyse participants who are healthy or only have one condition for the problem of multimorbidity. We showed that graphical lasso alleviated this step as the graph learning technique was able to produce reliable multimorbidity graphs with high predictive value even when healthy individuals or people with only one condition were considered. These graphbased models were able to predict with high accuracy conditions related to liver fibrosis such as ultrasound-based image patterns for fibrosed vessels (prominent pipe stems) and extensive liver fibrosis extending to the parenchyma and liver capsule as well as observations of severe cases of ascites and splenorenal shunts. Additionally, demonstrating the validity of the graph, the most common connections were between the liver patterns of prominent peripheral rings and prominent pipe stems which are two different cross-section views of mildly fibrosed vessels. Critically, we find that the predominant method of understanding multimorbidity that uses unadjusted observed co-occurrence for multimorbidity fails to establish relevant thresholds and does not allow for graph quality evaluation with no evidence for utility in predictive tasks related to hepatosplenic conditions.

## Implications of all available evidence

At worst, non-thresholded and unvalidated graphs could not only present irrelevant associations between conditions, but also could include artefacts specific to the dataset used that are not suitable for clinical decision-making and not generalisable to other populations. We showed that achieving sparsity in hepatosplenic multimorbidity graphs is critical for clinical decision-making. Sparsity enables a focus on condition inter-dependencies that co-occur with higher confidence in order to avoid misdiagnosis or mistreatment and to save limited resources needed for case management for already constrained health systems. The thresholded and validated hepatosplenic multimorbidity graphs presented in this study, if tested elsewhere, could provide clinical insight into the inter-dependencies of complex conditions in low-income settings, and can be used to guide screening and diagnostic strategies via ultrasound. Most importantly, the multimorbidity modelling pipeline proposed here is anticipated to be generalisable to systems beyond hepatosplenic multimorbidity and may guide more broadly the choice of multimorbidity outcomes.

## Notes

### Competing Interest Statement

The authors have declared no competing interest.

## References

1 Nguyen H, Manolova G, Daskalopoulou C, Vitoratou S, Prince M, Prina AM. Prevalence of multimorbidity in community settings: A systematic review and metaanalysis of observational studies. Journal of comorbidity. 2019;9:2235042X19870934.

2 Barnett K, Mercer SW, Norbury M, Watt G, Wyke S, Guthrie B. Epidemiology of multimorbidity and implications for health care, research, and medical education: a cross-sectional study. The Lancet. 2012;380(9836):37-43.

3 Chami GF, Kabatereine NB, Tukahebwa EM, Dunne DW. Precision global health and comorbidity: a population-based study of 16 357 people in rural Uganda. Journal of The Royal Society Interface. 2018;15(147):20180248.

4 Academy of Medical Sciences RU. Multimorbidity: a priority for global health research. Academy of medical sciences; 2018.

5 Schäfer I, von Leitner EC, Schön G, Koller D, Hansen H, Kolonko T, et al. Multimorbidity patterns in the elderly: a new approach of disease clustering identifies complex interrelations between chronic conditions. PloS one. 2010;5(12):e15941.

6 Xu Z, Zhang Q, Yip PSF. Predicting post-discharge self-harm incidents using disease comorbidity networks: A retrospective machine learning study. Journal of affective disorders. 2020;277:402–9.

7 Woodman RJ, Koczwara B, Mangoni AA. Applying precision medicine principles to the management of multimorbidity: the utility of comorbidity networks, graph machine learning, and knowledge graphs. Frontiers in Medicine. 2024;10:1302844.

8 Wang T, Bendayan R, Msosa Y, Pritchard M, Roberts A, Stewart R, et al. Patient-centric characterization of multimorbidity trajectories in patients with severe mental illnesses: A temporal bipartite network modeling approach. Journal of Biomedical Informatics. 2022;127:104010.

9 Lee YJ, Boyd AD, Gardeux V, Kenost C, Saner D, Li H, et al. COPD hospitalization risk increased with distinct patterns of multiple systems comorbidities unveiled by network modeling. In: AMIA Annual Symposium Proceedings. vol. 2014. American Medical Informatics Association; 2014. p. 855.

10 Park J, Lee DS, Christakis NA, Barabási AL. The impact of cellular networks on disease comorbidity. Molecular systems biology. 2009;5(1):262.

11 Xu H, Moni MA, Liò P. Network regularised cox regression and multiplex network models to predict disease comorbidities and survival of cancer. Computational biology and chemistry. 2015;59:15–31.

12 Nam Y, Jung SH, Verma A, Sriram V, Won HH, Yun JS, et al. netCRS: Networkbased comorbidity risk score for prediction of myocardial infarction using biobank-scaled PheWAS data. In: PACIFIC SYMPOSIUM ON BIOCOMPUTING 2022. World Scientific; 2021. p. 325–36.

13 Zhao B, Huepenbecker S, Zhu G, Rajan SS, Fujimoto K, Luo X. Comorbidity network analysis using graphical models for electronic health records. Frontiers in Big Data. 2023;6.

14 Cao Y, Raoof M, Szabo E, Ottosson J, Näslund I. Using Bayesian networks to predict long-term health-related quality of life and comorbidity after bariatric surgery: A study based on the Scandinavian obesity surgery registry. Journal of clinical medicine. 2020;9(6):1895.

15 Lappenschaar M, Hommersom A, Lucas PJ, Lagro J, Visscher S, Korevaar JC, et al. Multilevel temporal Bayesian networks can model longitudinal change in multimorbidity. Journal of clinical epidemiology. 2013;66(12):1405–16.

16 Nuffield Department of Population Health UoO. SchistoTrack: a prospective multimorbidity cohort.; Date accessed: 2024-08-20. Accessed: 2024-08-06. Available from:.

17 Anjorin S, Nabatte B, Mpooya S, Tinkitina B, Opio CK, Kabatereine NB, et al. The epidemiology of periportal fibrosis and relevance of current Schistosoma mansoni infection: a population-based, cross-sectional study. medRxiv. 2023:2023–09.

18 Woodman RJ, Koczwara B, Mangoni AA. Applying precision medicine principles to the management of multimorbidity: the utility of comorbidity networks, graph machine learning, and knowledge graphs. Frontiers in Medicine. 2023;10.

19 Peel L, Peixoto TP, De Domenico M. Statistical inference links data and theory in network science. Nature Communications. 2022;13(1):6794.

20 Friedman J, Hastie T, Tibshirani R. Sparse inverse covariance estimation with the graphical lasso. Biostatistics. 2008;9(3):432–41.

21 Borgwardt K, Ghisu E, Llinares-López F, O’Bray L, Rieck B, et al. Graph kernels: State-of-the-art and future challenges. Foundations and Trends® in Machine Learning. 2020;13(5-6):531–712.

22 Shervashidze N, Schweitzer P, Van Leeuwen EJ, Mehlhorn K, Borgwardt KM. Weisfeiler-lehman graph kernels. Journal of Machine Learning Research. 2011;12(9).

23 Kriege N, Mutzel P. Subgraph matching kernels for attributed graphs. arXiv preprint arXiv:12066483. 2012.

24 Hido S, Kashima H. A linear-time graph kernel. In: 2009 Ninth IEEE International Conference on Data Mining. IEEE; 2009. p. 179–88.

25 Kipf TN, Welling M. Semi-supervised classification with graph convolutional networks. arXiv preprint arXiv:160902907. 2016.

26 Veličković P, Cucurull G, Casanova A, Romero A, Lio P, Bengio Y. Graph attention networks. arXiv preprint arXiv:171010903. 2017.

27 Hamilton W, Ying Z, Leskovec J. Inductive representation learning on large graphs. Advances in neural information processing systems. 2017;30.

28 Munikoti S, Agarwal D, Das L, Halappanavar M, Natarajan B. Challenges and Opportunities in Deep Reinforcement Learning with Graph Neural Networks: A Comprehensive review of Algorithms and Applications. IEEE transactions on neural networks and learning systems. 2022;PP. Available from:.

29 Dong G, Zhang ZC, Feng J, Zhao XM. MorbidGCN: prediction of multimorbidity with a graph convolutional network based on integration of population phenotypes and disease network. Briefings in Bioinformatics. 2022;23(4):bbac255.

30 Zhi YC, Ng YC, Dong X. Gaussian processes on graphs via spectral kernel learning. IEEE Transactions on Signal and Information Processing over Networks. 2023;9:304-14.

