## Appendix for "Multimorbidity representation via graph learning: A population-based study on hepatosplenic conditions in schistosomiasis-endemic areas of rural Uganda"

#### Hepatosplenic condition definitions

Ultrasound assessments were conducted on site by four local sonographers following procedures described in Anjorin et al.[1] and the Niamey Protocol,[2] and later reviewed through random assignment of images and videos to a different sonographer among the original four sonographers for quality assurance. Image patterns representative of periportal and portal fibrosis were identified and classified as demonstrable fibrosis but unclear of pattern (B), feather streaks (small echogenic lines, B0), flying saucers or starry sky (small diffuse echogenic nodules, B1), spider thickening (B2), prominent peripheral rings (C1), prompinent pipe stems (C2), extensive thickening 'ruff' around the portal bifurcation (D), occluded fibrosed vessels extending into the liver parenchyma (E), and extensive fibrosis extending to the liver capsule edge (F). The following abnormalities also were diagnosed in each individual including cirrhosis-like livers, fatty-like livers, chronic hepatitis-B-like livers (hepatitis B only as there was hepatitis C in these areas of Uganda), and fatty-like livers; all described in detail in the supplementary methods of Anjorin et al.[1] Evidence of polycystic kidney disease, liver cysts, situs inversus, and any other abnormalities also were recorded. Liver surface irregularities were classified, if present, as slight (serrated surface) or (gross) undulating surface. The caudal liver edge was considered abnormal if it was rounded instead of sharp.

For the left and right liver lobes, spleen, and portal vein diameter, we measured organometry against an internal healthy reference population standardised by height. The internal reference population excluded pregnant women, individuals with focal or diffuse liver fibrosis patterns, and individuals who were reported to have had their spleens removed. The reference population was then divided by gender and into height deciles, and participants were evaluated in comparison to the mean of their division where one standard deviation away from the mean was moderately morbid and two or more standard deviations was severely morbid. Above or below the mean were interpreted as enlarged or restricted/shrunken, respectively. The left liver lobe length was measured from the left parasternal line with the abdominal aorta as reference. The right liver lobe length was measured from the midclavicular line with the gall bladder as the reference. Liver lobe sizes were restricted to a possible range of 2-25cm. The upper and lower tips of the spleen length also were measured at the two poles with a requirement that the central line cross the splenic hilum. Two measurements of both the inner diameter of the main portal vein near its entry to liver were taken. The mean was taken and compared to the reference population in the same manner as the liver and spleen sizes with restricted possible range of 0·1-0·6cm.

Any portosystemic collaterals of the liver also were recorded, including splenic varices, gastro-oesophageal varices, pancreaticoduodenal varices, an entirely recanalized paraumbilical vein greater or equal to 3mm, splenorenal shunts, and any other collaterals. Ascites were diagnosed where visible regardless of the extent of abdominal cavity covered. Complications of the gall bladder were recorded such as if it was not visible on ultrasound, blocked by a stone, or collapsed. Gall bladder walls were measured and thickness of greater than 0·3cm was noted as thickened (approximately the top 10% of values; 87·6 percentile to be exact) where the observed range was 0·05-1·87cm.

(a)

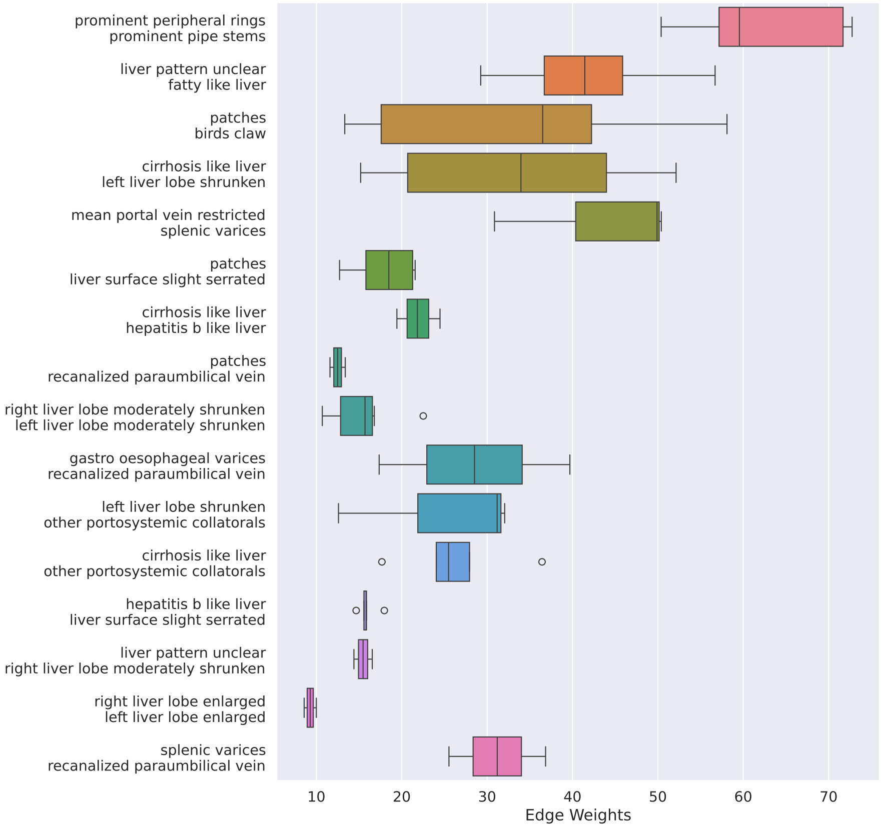


(b)

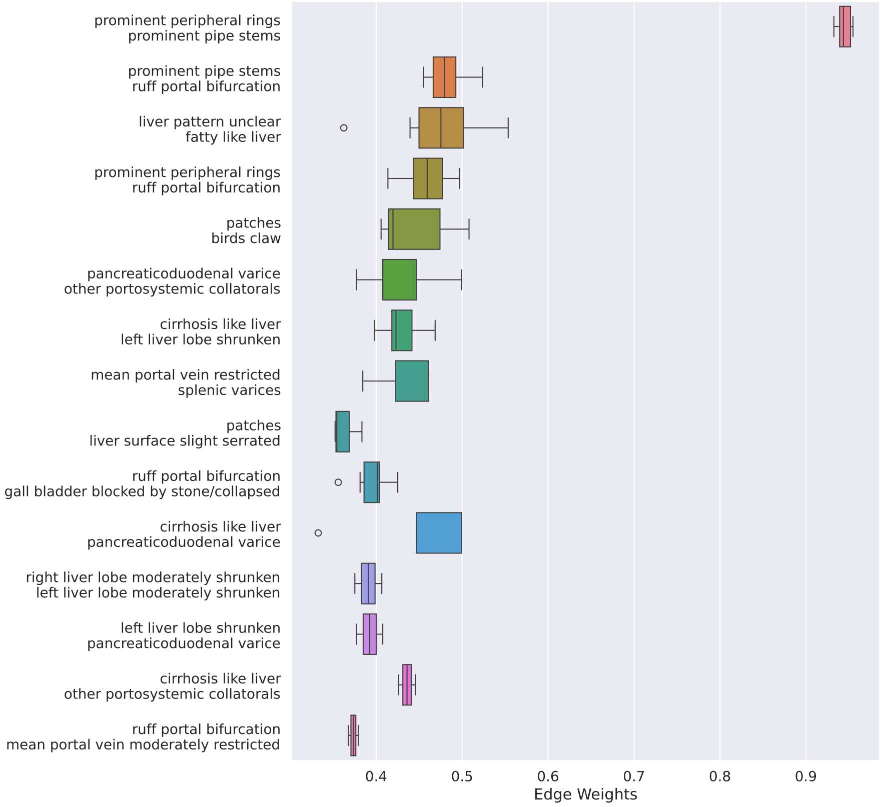


Figure S1: Distribution of top 2% most occurring edges learned on the mixed population. Edges are found from 500 samples, (a) graphical lasso (b) signed distance correlation.

(a)

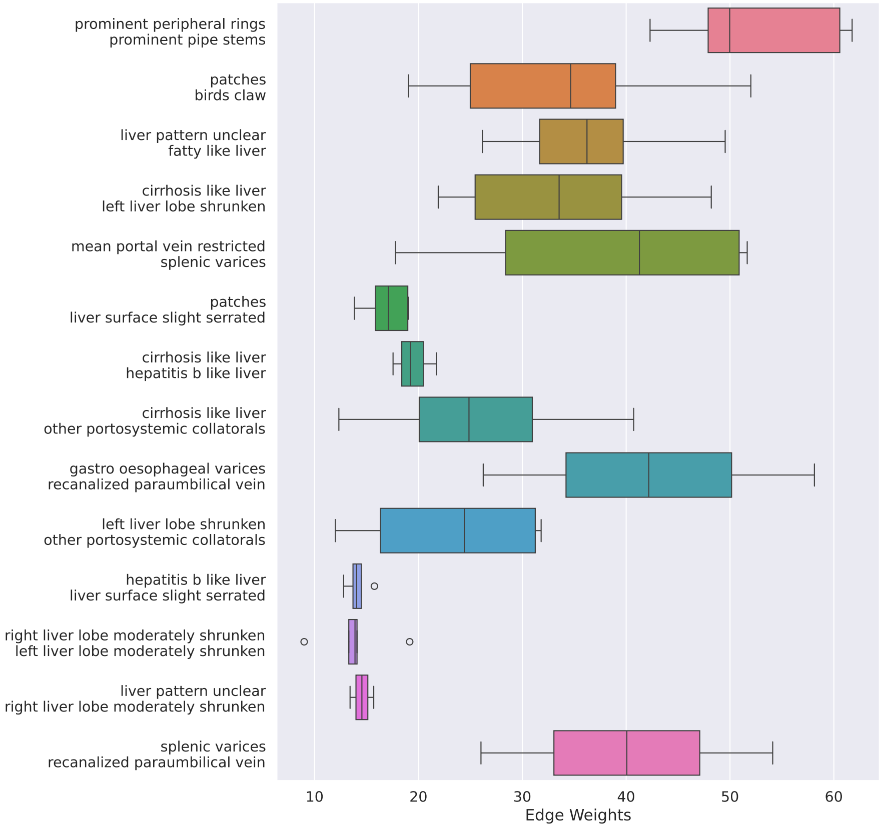


(b)

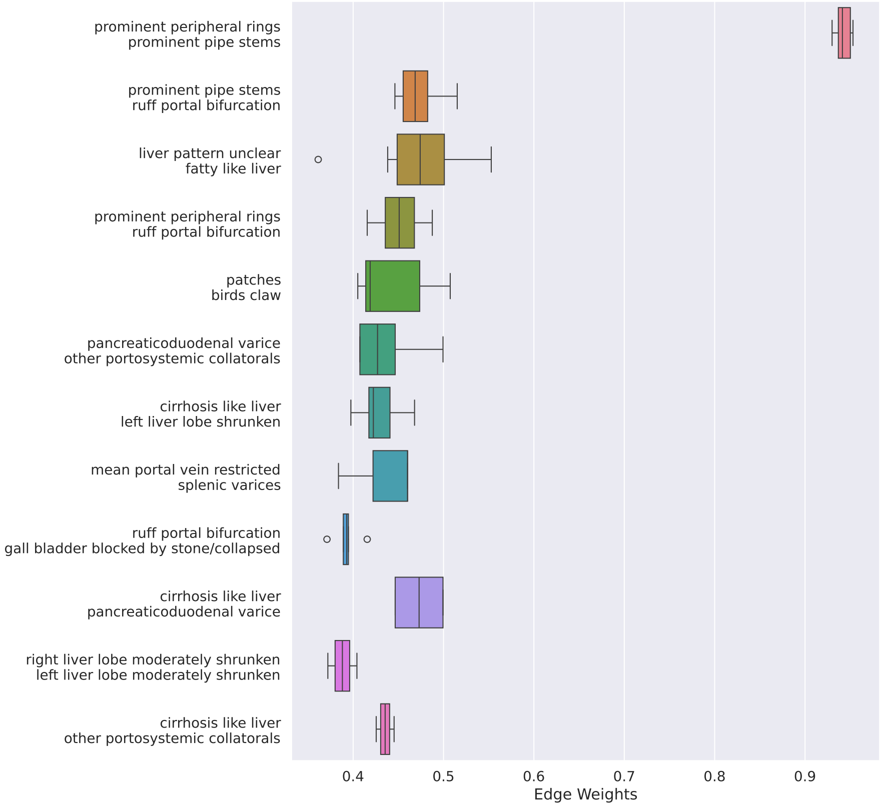


Figure S2: Distribution of top 2% most occurring edges learned on the morbid population.
Edges are found from 500 samples, (a) graphical lasso (b) signed distance correlation.

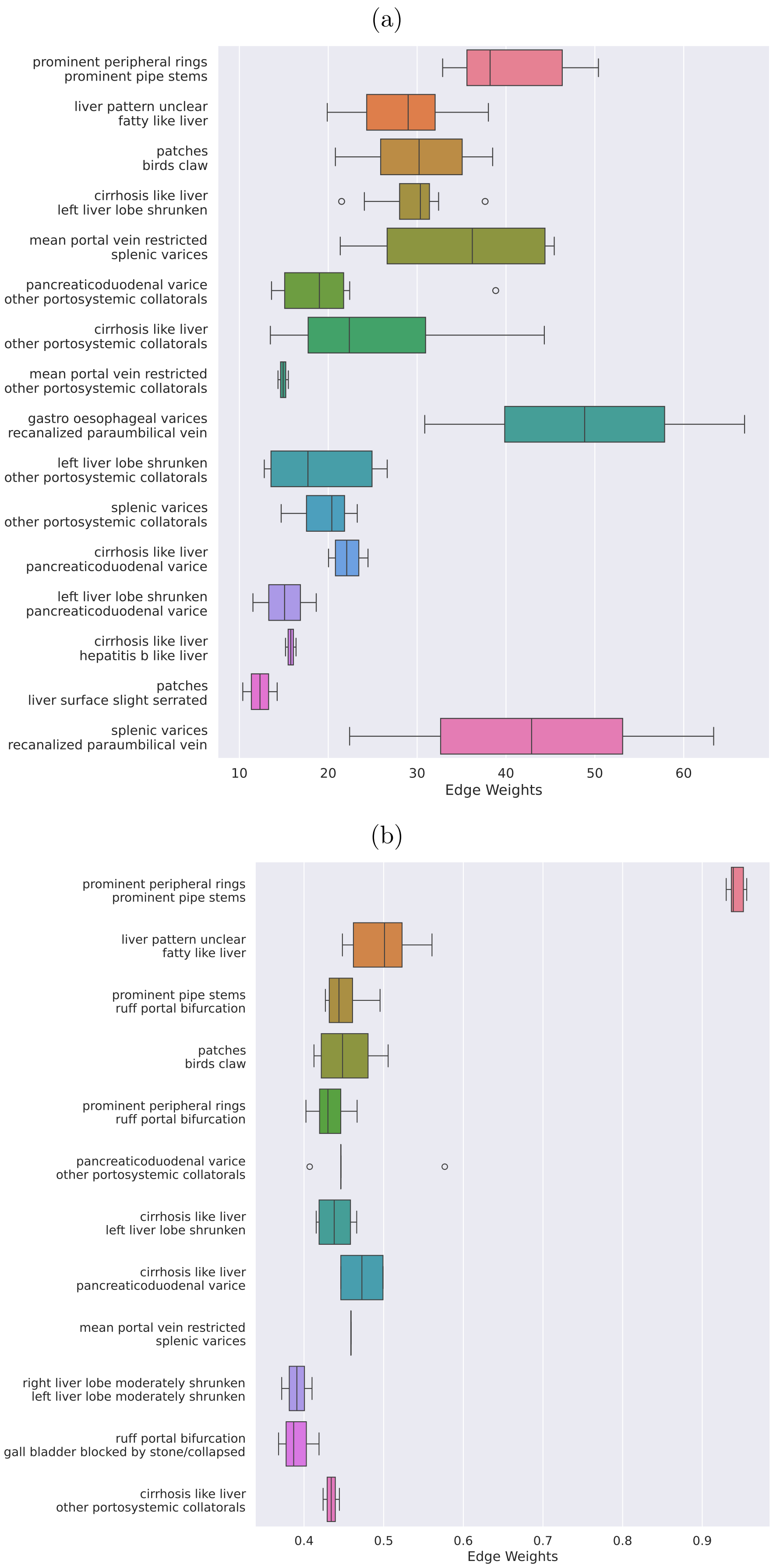


Figure S3: Distribution of top 2% most occurring edges learned on the multimorbid population.
Edges are found from 500 samples, (a) graphical lasso (b) signed distance correlation.

#### Graph Kernels

The Weisfeiler-Lehman kernel[3] is an adaptation of the Weisfeiler-Lehman test for graph isomorphism[4]; the kernel compares the similarity of graphs by recursively refining node labels based on their 1-hop neighborhoods. Subgraph matching kernel[5] computes and compares subgraphs within the two input graphs that correspond to the maximum cliques in the product graph. Neighborhood hash kernel[6] constructs node embeddings by converting each node label value into binary, and compute new labels by combining the 1-hop neighbours labels through binary operators ($XOR$ and $ROT$).

#### Graph Neural Networks

Graph convolutional networks (GCNs):[7] layer $l$ updates the embeddings for node $i$ as $\mathbf{x}_i^{l+1} = \mathbf{W}^\top \sum_{j \in \mathcal{N}(i) \cup i} \mathbf{x}_j^l \hat{\mathbf{A}}_{ij} / \sqrt{\hat{d}_i \hat{d}_j}$ for trainable neural network weights $\mathbf{W}$, adjacency matrix with self loop $\hat{\mathbf{A}}$, and $\hat{d}_i, \hat{d}_j$ are the degrees of $\hat{\mathbf{A}}$. This convolution has the effect of averaging node embeddings with their neighbours in each layer, inducing the graph information by making connected nodes more similar.

Graph attention networks (GATs):[8] layer $l$ updates the node embeddings as $\mathbf{x}_i^{l+1} = \mathbf{W}^\top \sum_{j \in \mathcal{N}(i) \cup i} \mathbf{x}_j^l \alpha_{ij} \hat{\mathbf{A}}_{ij} / \sqrt{\hat{d}_i \hat{d}_j}$. This builds on GCN with the addition of trainable attention coefficients $\alpha_{ij}$ to weigh the influence of each neighbouring nodes.

Sample and aggregate (GraphSAGE):[9] instead of using the full neighbourhood, a random fixed size sample (with replacement) of nodes from $\mathcal{N}(i)$ is used to aggregate for each node. The stochasticity of the neighbour samples allow for the model to translate better to unseen data. The aggregate function is kept as the mean like previous models.

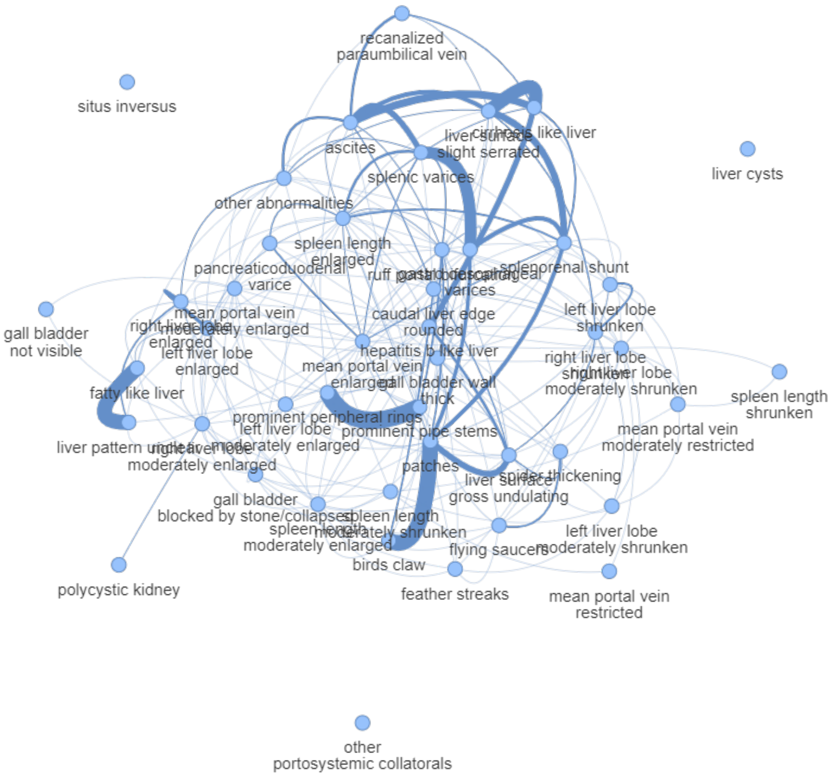


Figure S4: Initial graphical lasso graph without any thresholding.

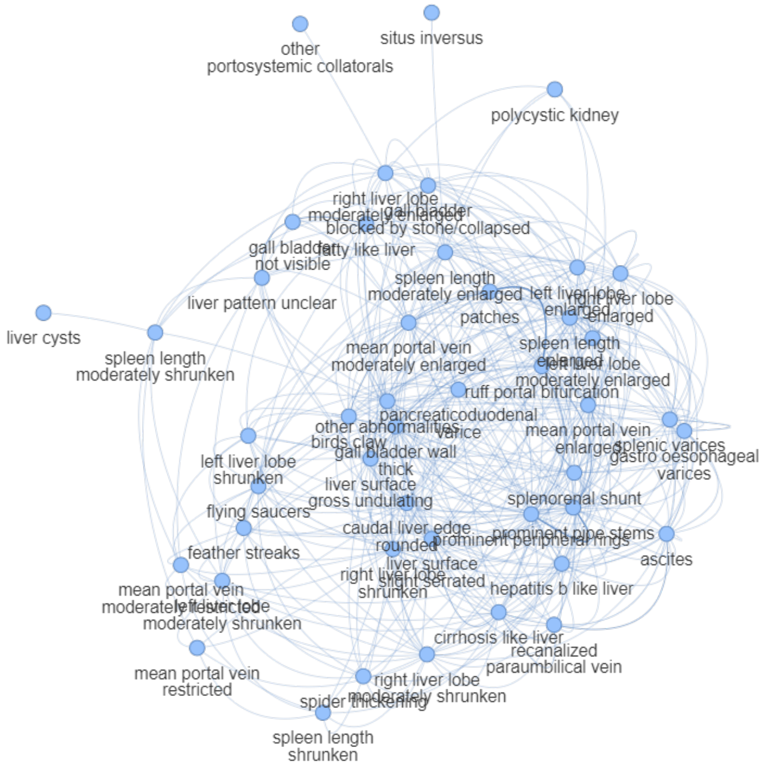


Figure S5: Initial signed distance correlation graph without any thresholding.

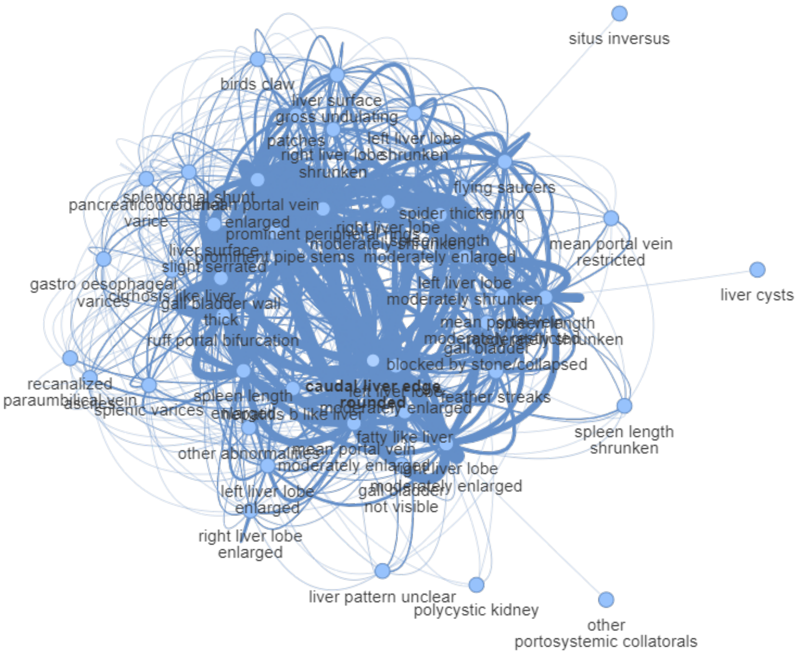


Figure S6: Co-occurrence graph inherently without any thresholding.

| | Graphical Lasso | Signed Distance Correlation | Co-Occurrence |
|---|---|---|---|
| Number of Edges | 221·51 ± 9·18 | 422·56 ± 13·83 | 545·20 ± 14·08 |
| Average Degree | 9·84 ± 0·41 | 18·78 ± 0·61 | 24·23 ± 0·63 |
| Average Clustering Coefficient | 0·41 ± 0·02 | 0·61 ± 0·01 | 0·75 ± 0·01 |
| Diameter | 3·77 ± 0·45 | 3·30 ± 0·47 | 3·15 ± 0·36 |
| Edge Density | 0·22 ± 0·01 | 0·42 ± 0·01 | 0·55 ± 0·01 |
| Largest Component Size | 42·00 ± 0·71 | 45·00 ± 0·00 | 45·00 ± 0·00 |
| Number of Isolated Nodes | 3·00 ± 0·71 | 0·00 ± 0·00 | 0·00 ± 0·00 |
| Assortativity | -0.05 ± 0.04 | 0.02 ± 0.04 | -0.21 ± 0.02 |
| Largest Component Spectral Gap | 0.83 ± 0.56 | 0.05 ± 0.03 | 1.00 ± 0.00 |
| Degree Distribution | Power law | None | Power law |

Table S1: Global statistics for unthresholded graphs averaged over the 500 samples. Degree distribution of "None" indicated that the graphs did not fit any of exponential, power law, or log-normal distributions.

(a)

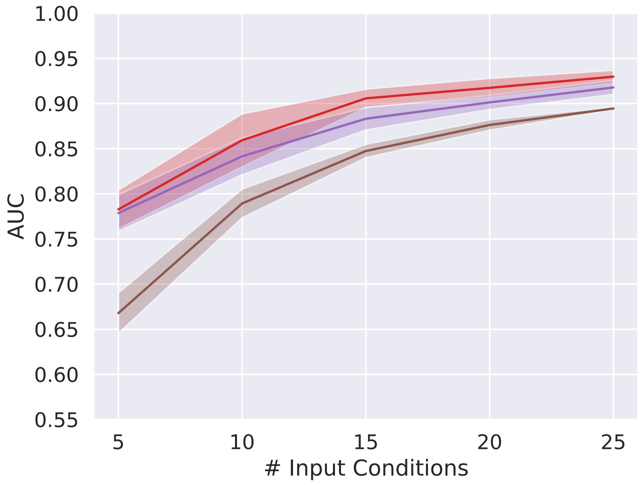


(b)

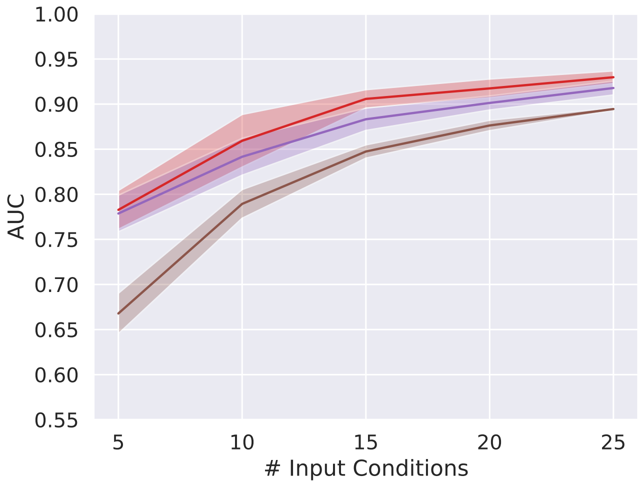


(c)

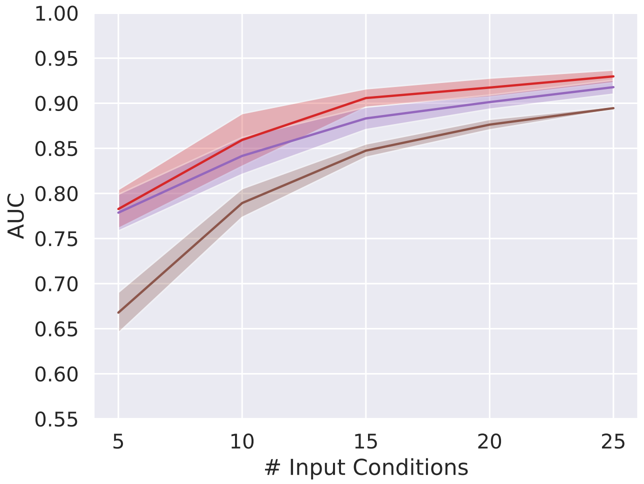


Figure S7: Multimorbidity prediction on unobserved conditions only with varying number of inputs.
(a) GCN, (b) GAT, & (c) GraphSAGE, the model uses graphs with optimal thresholds, Red: graphical lasso 50·16%, Purple: signed distance correlation 64·46%, Brown: Co-occurrence 0%. All AUCs were averaged over 10 training-testing splits.

#### Further analyses for GNN predictions

In testing the GNNs over varying thresholds instead of just the optimal threshold chosen by the graph kernels, GCN (Appendix Fig. S8a) and GAT (Appendix Fig. S8b) predictions with graphical lasso and signed distance correlation generally did not have clear peak performances with these instead falling in the large range of thresholds 30-65%, although this range included the threshold captured by the graph kernels. The co-occurrence graph; however, led to noticeably different behaviours, generally lacking any clear pattern with respect to edge removal. For GraphSAGE (Fig. S8c), the patterns from graphical lasso and signed distance correlation were similar with the removal of edges generally leading to deteriorating performances as would be expected in the GNN made use of the graph structure.

Appendix Fig. S9 - S18 shows more comprehensive comparisons of the GNNs on the three population splits. $m$ was set to five conditions to maximise the difference between the GNNs.Across the three populations, there were only minor differences in the performance between all GNN models. On GCN and GAT, graphical lasso and signed distance correlation were again quite consistent in showing the best performances when the thresholds were again between 30-65%. The co-occurrence graph led to much more erratic behaviours, in GCN it did not improve with respect to any threshold, but with GraphSAGE it needed close to full removal of the graph to see a performance drop, again indicating most of the graph structure made little difference in aiding the model performance.

(a)

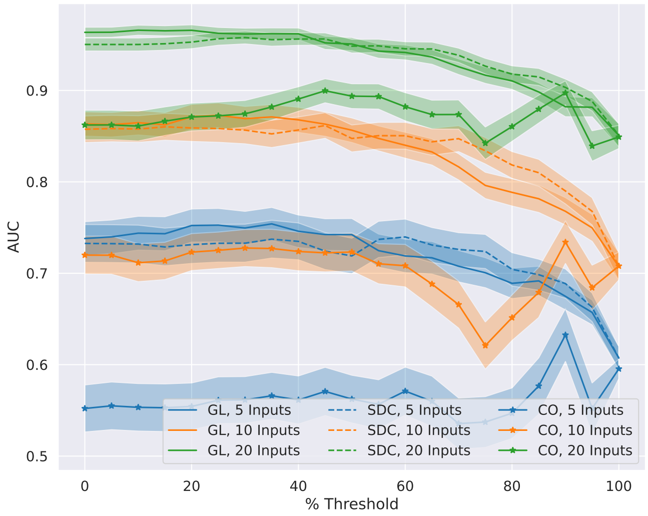


(b)

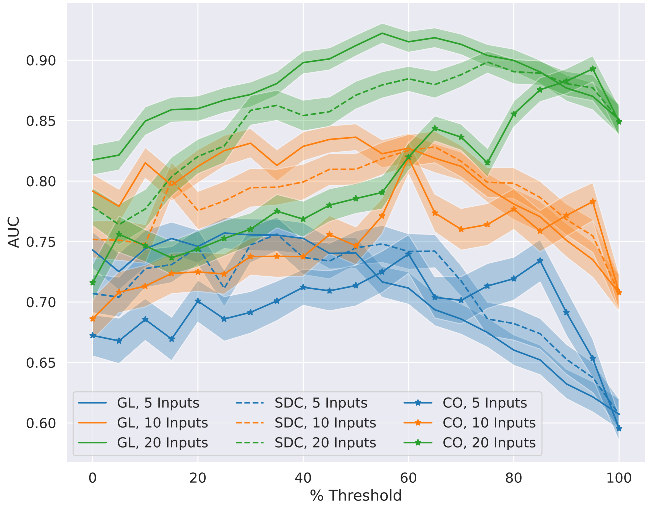


(c)

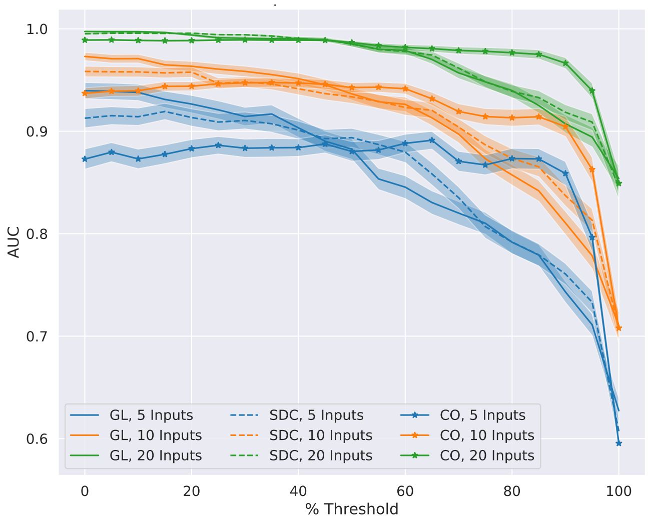


Figure S8: Multimorbidity prediction with percentage thresholding and varying numbers of input conditions.
(a) GCN, (b) GAT, & (c) GraphSAGE, $X\%$ of the smallest edges were removed from the graph before being passed into the GNN.

(a)

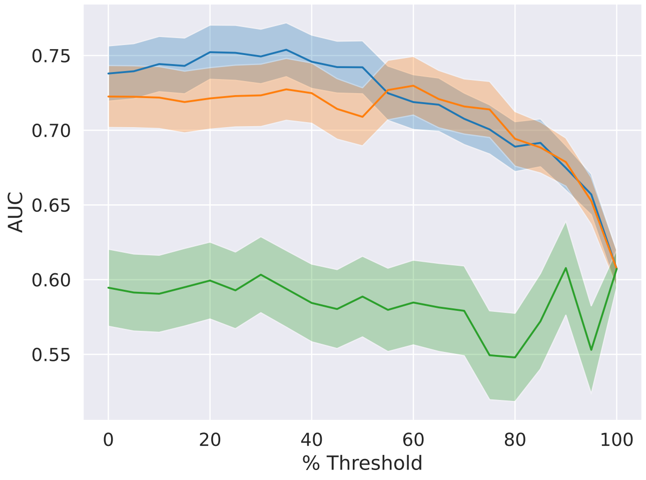


(b)

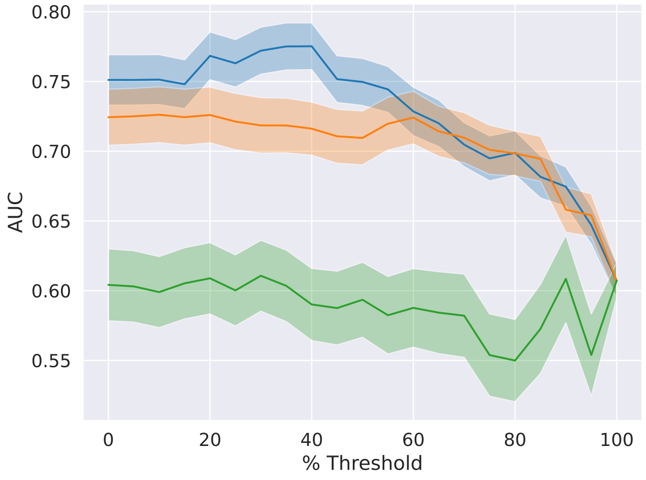


(c)

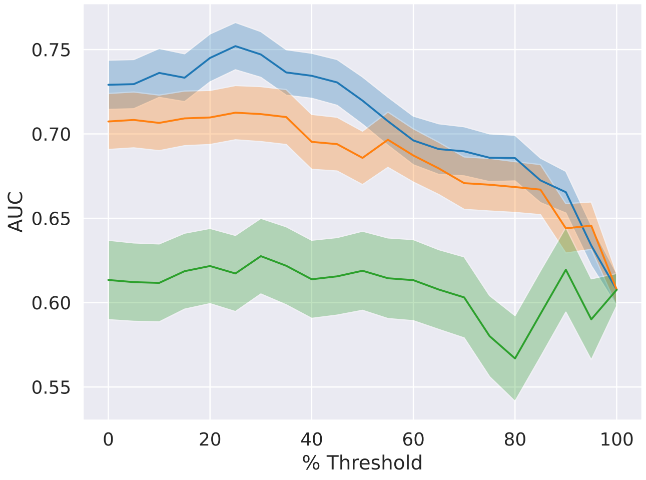


Figure S9: GCN Multimorbidity prediction on various populations.
In each plot, the graph was learned on the same population used to train the GNN. Thresholding removes $X\%$ of the smallest edge weights. Blue: graphical lasso, Orange: signed distance correlation, Green: co-occurrence. (a) full population (d) morbid population (g) multimorbid population

(a)

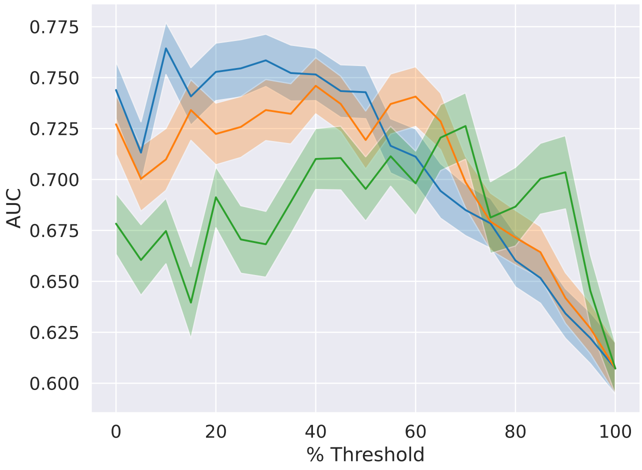


(b)

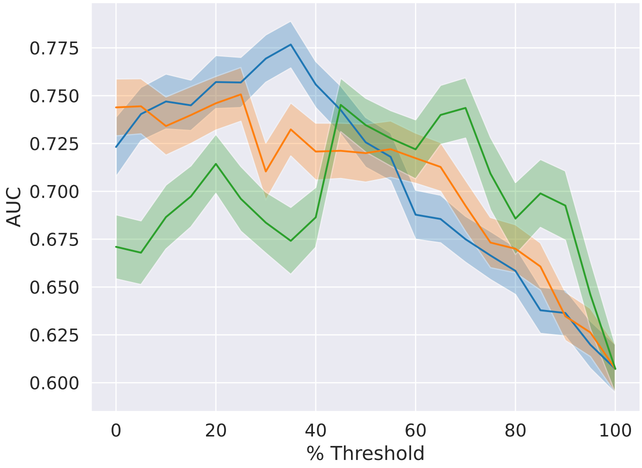


(c)

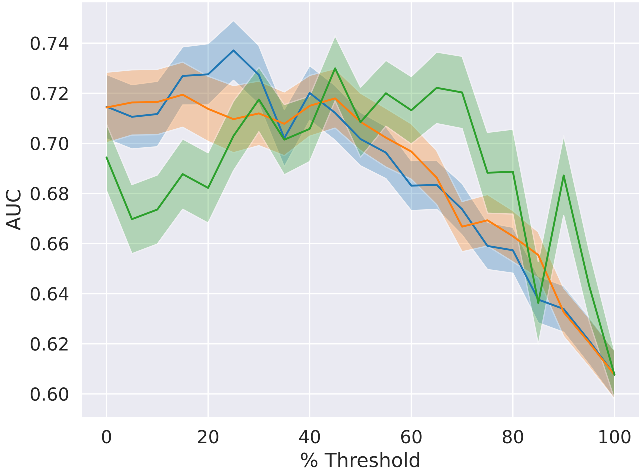


Figure S10: GAT Multimorbidity prediction on various populations.
In each plot, the graph was learned on the same population used to train the GNN. Thresholding removes $X\%$ of the smallest edge weights. Blue: graphical lasso, Orange: signed distance correlation, Green: co-occurrence. (a) full population (d) morbid population (g) multimorbid population

(a)

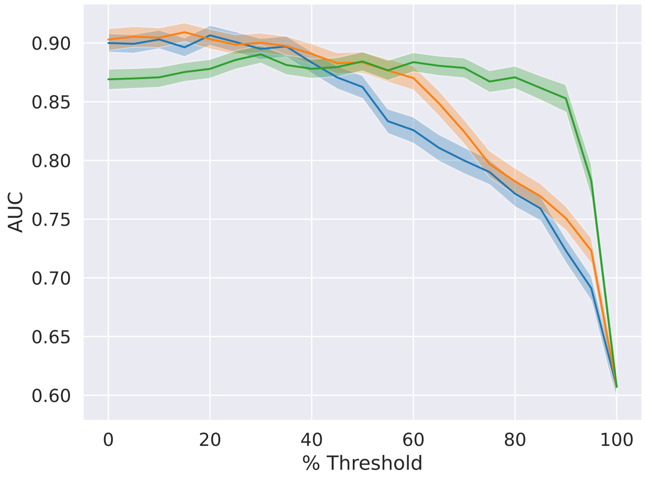


(b)

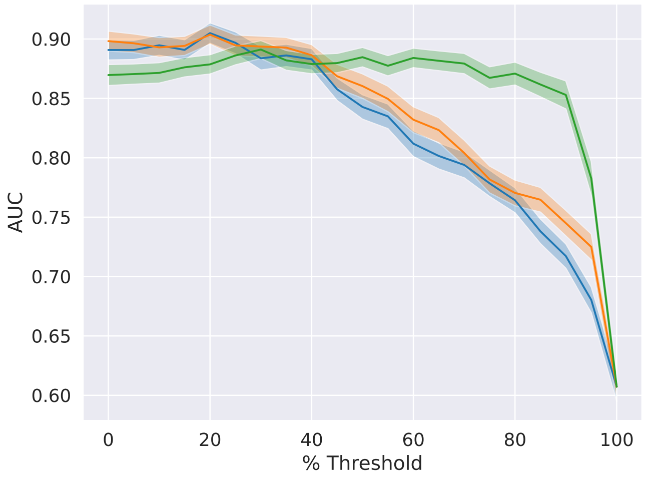


(c)

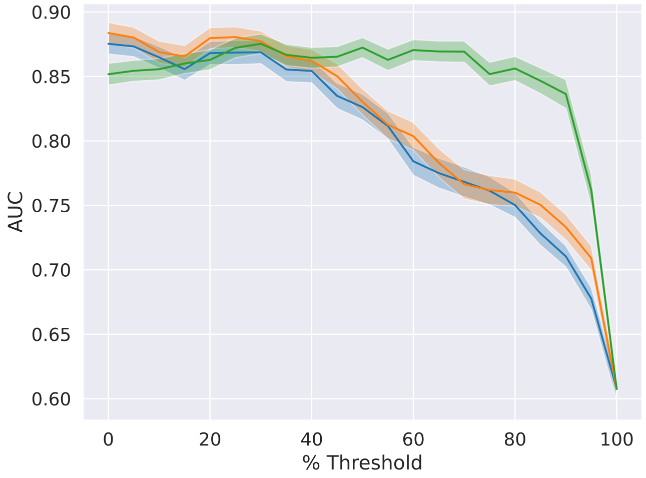


Figure S11: GraphSAGE Multimorbidity prediction on various populations.
In each plot, the graph was learned on the same population used to train the GNN. Thresholding removes $X\%$ of the smallest edge weights. Blue: graphical lasso, Orange: signed distance correlation, Green: co-occurrence. (a) full population (d) morbid population (g) multimorbid population

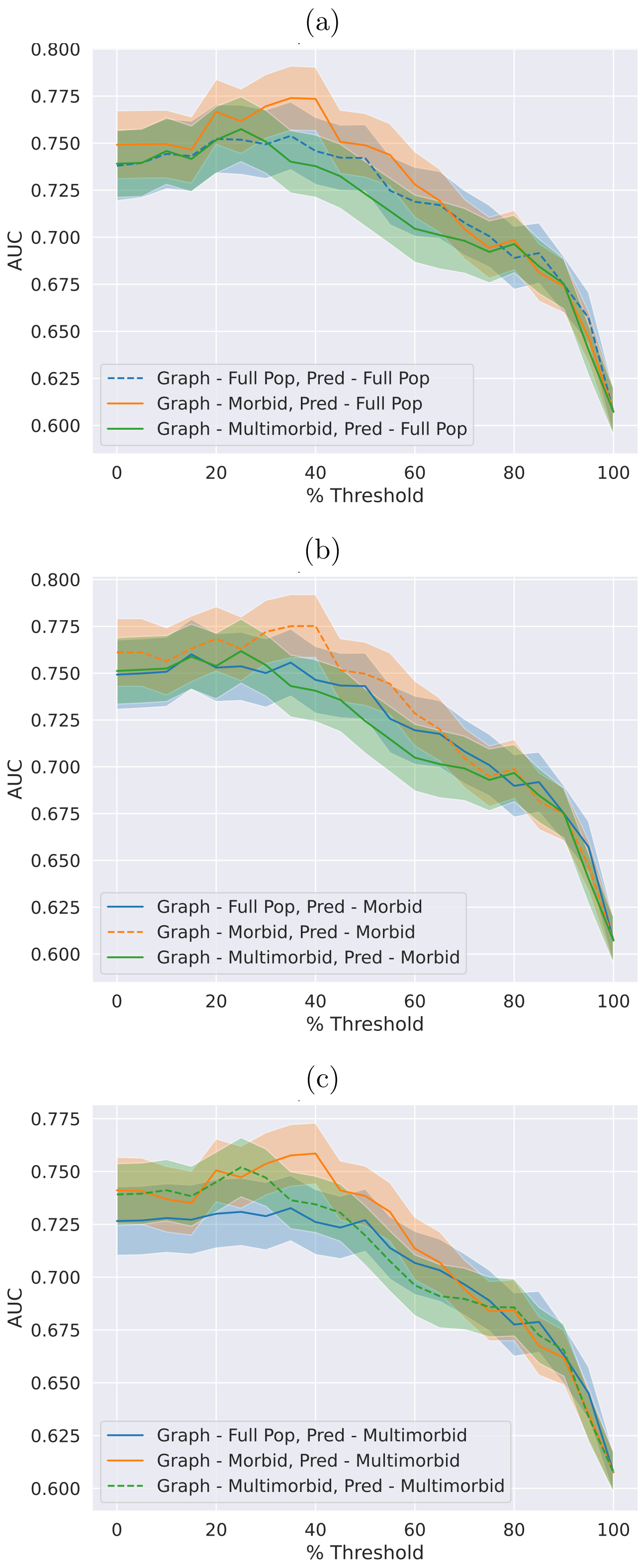


Figure S12: Cross-population prediction with GCN and graphical lasso.
An analysis is presented where different populations were used for graph learning and prediction. The dotted line indicates the graph was learned on the same population as prediction.

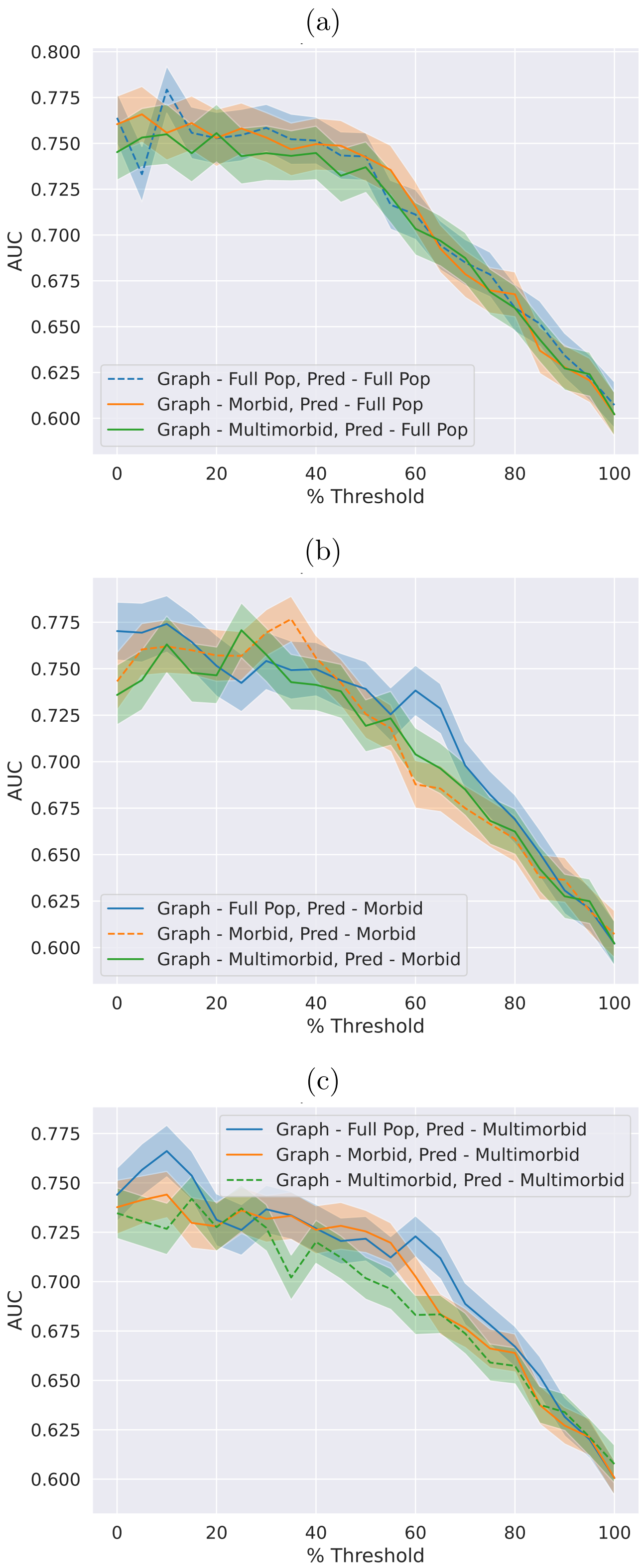


Figure S13: Cross-population prediction with GAT and graphical lasso.
An analysis of different populations for graph learning and prediction is presented. The dotted line indicates graph was learned on the same population as prediction.

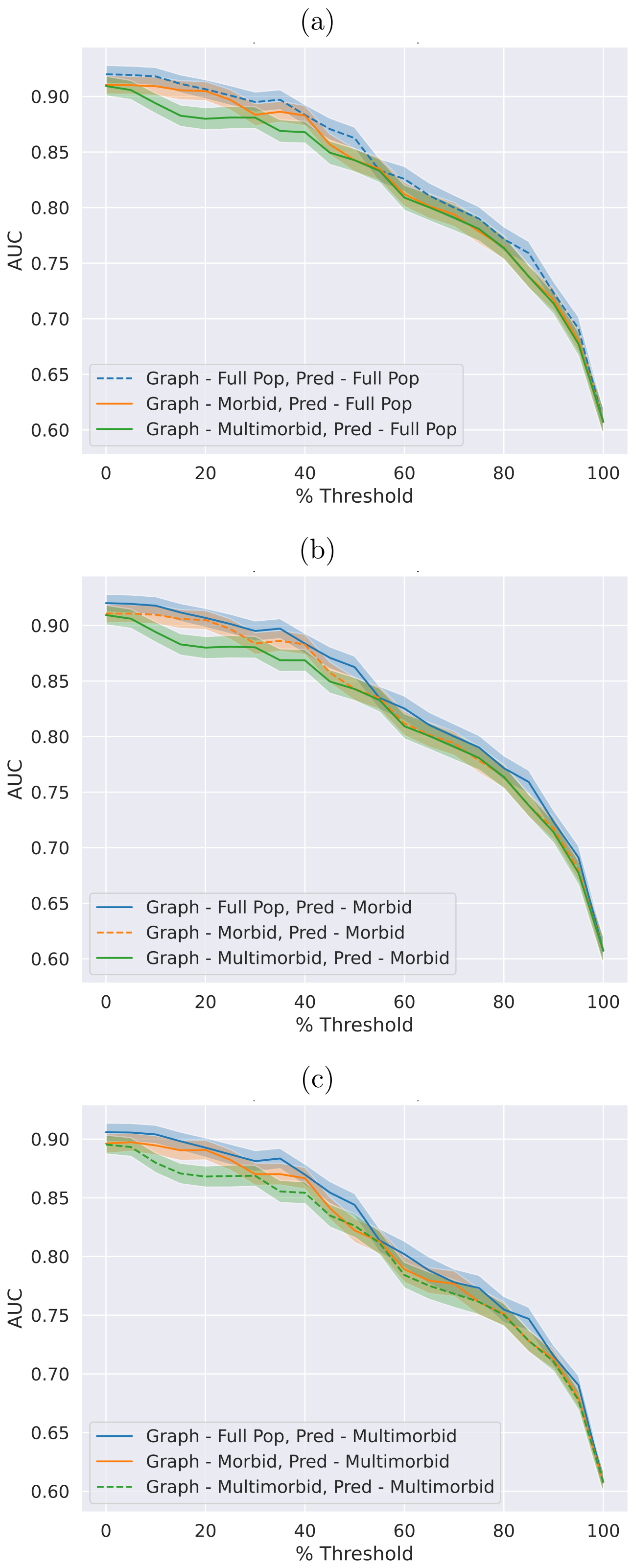


Figure S14: Cross-population prediction with GraphSAGE and graphical lasso. An analysis of different populations for graph learning and prediction is presented with the dotted line indicating that the graph was learned on the same population as prediction.

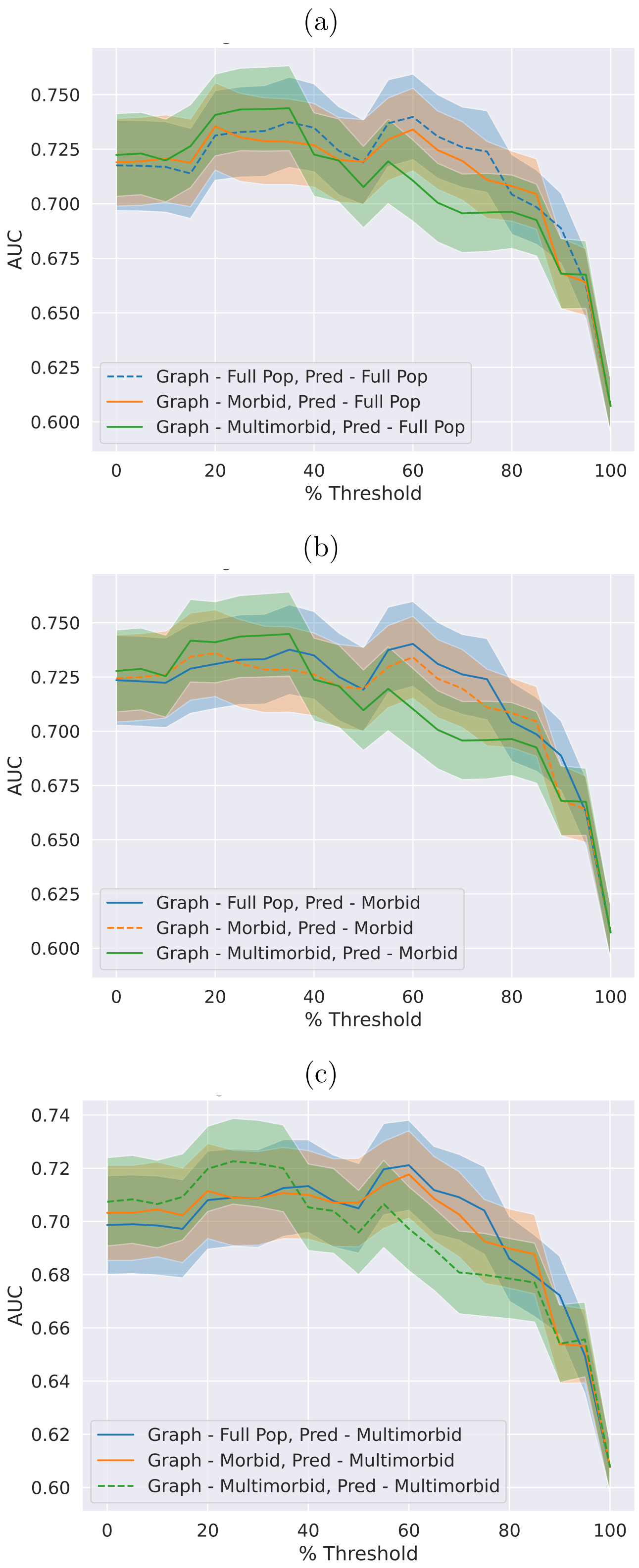


Figure S15: Cross-population prediction with GCN and signed distance correlation. An analysis of different populations for graph learning and prediction is presented. The dotted line indicates graph was learned on the same population as prediction.

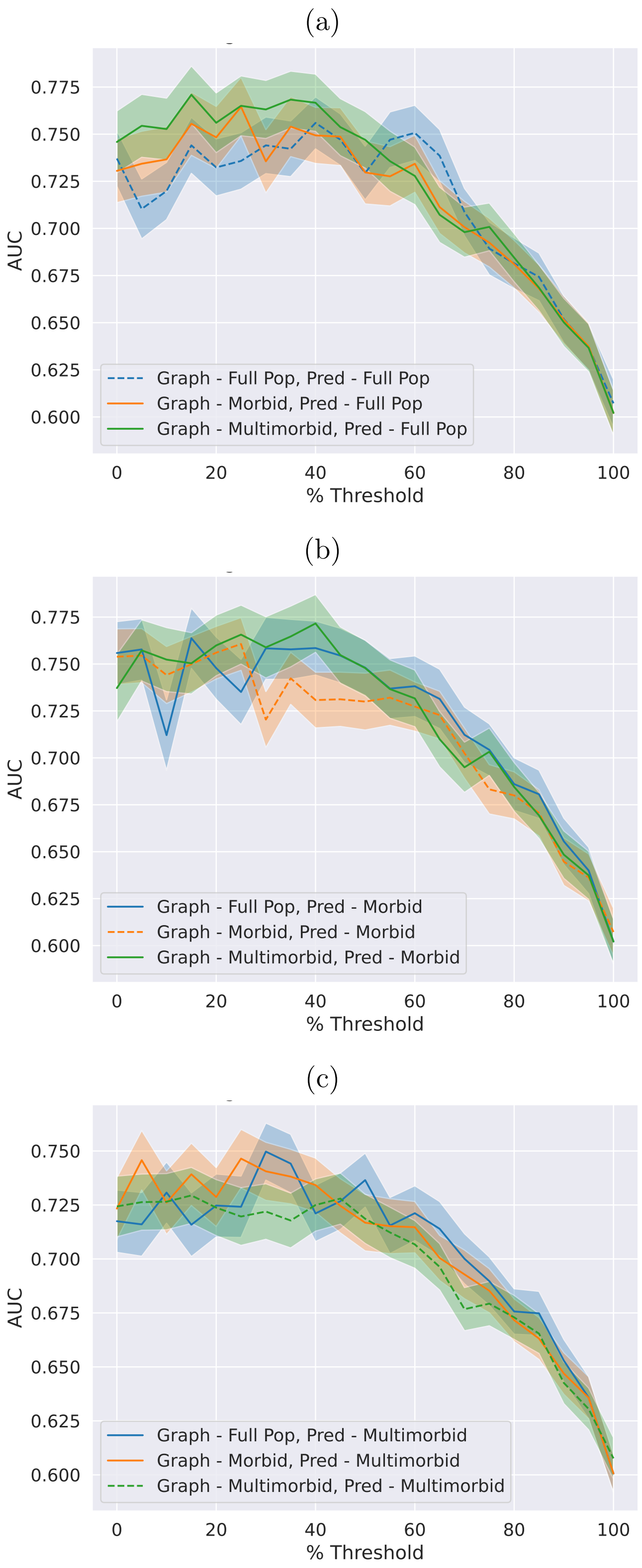


Figure S16: Cross-population prediction with GAT and signed distance correlation. An analysis of different populations for graph learning and prediction is presented. The dotted line indicates graph was learned on the same population as prediction.

(a)

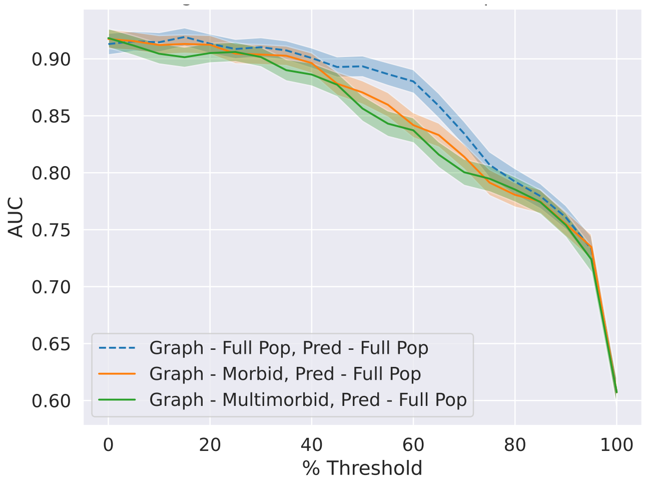


(b)

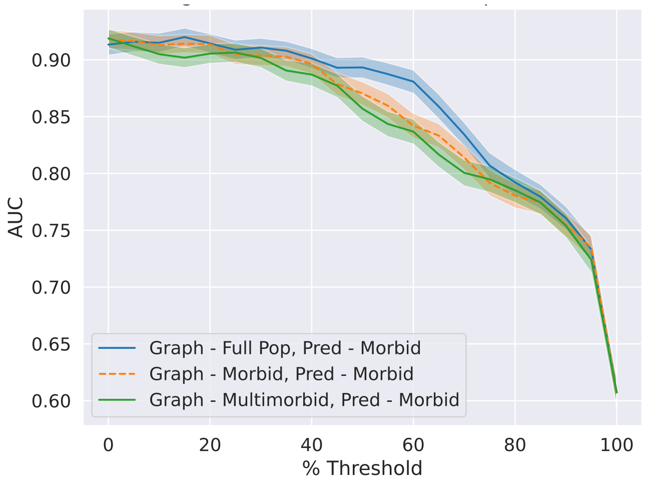


(c)

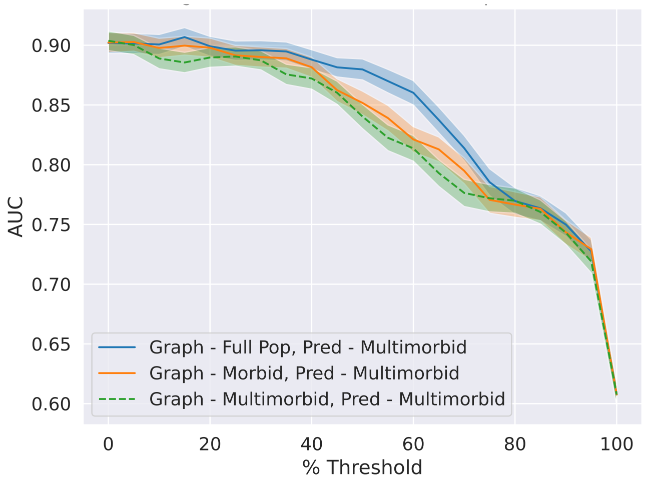


Figure S17: Cross-population prediction with GraphSAGE and signed distance correlation.
An analysis of different populations for graph learning and prediction is presented. The dotted line indicates graph was learned on the same population as prediction.

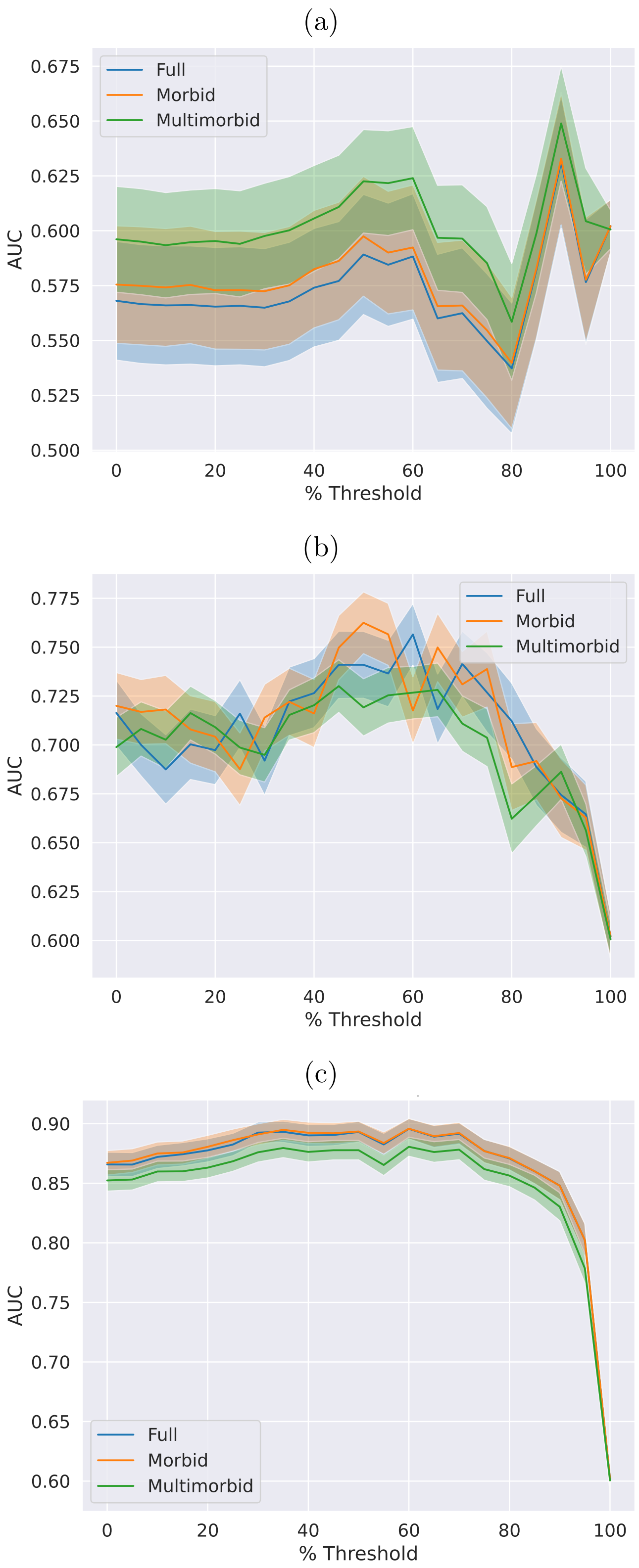


Figure S18: Cross-populations predictions with co-occurrence.
(a) GCN (b) GAT (c) GraphSAGE.

(a)

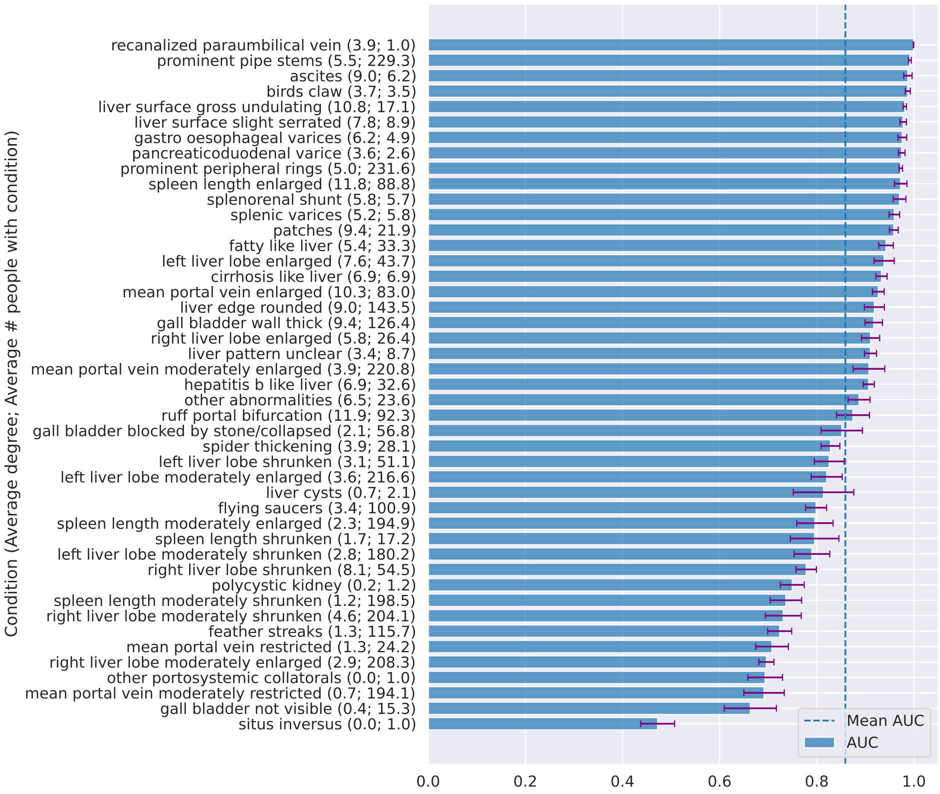


(b)

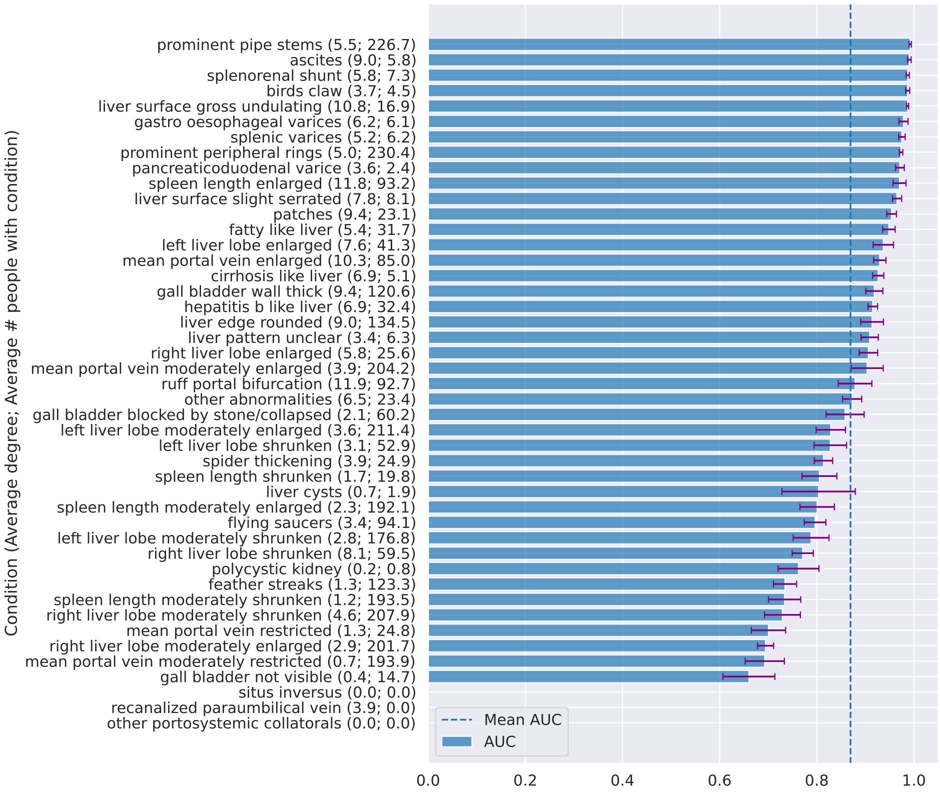


Figure S19: Performance on each condition ordered by AUC with GCN and graphical lasso.
This figure adds information to the results in Fig. 5. AUCs cannot be computed when the ground truth only consist of 1 class (when 0 people exhibited the condition) and therefore does not exist for some conditions on the test set. (a) training set (b) testing set.

(a)

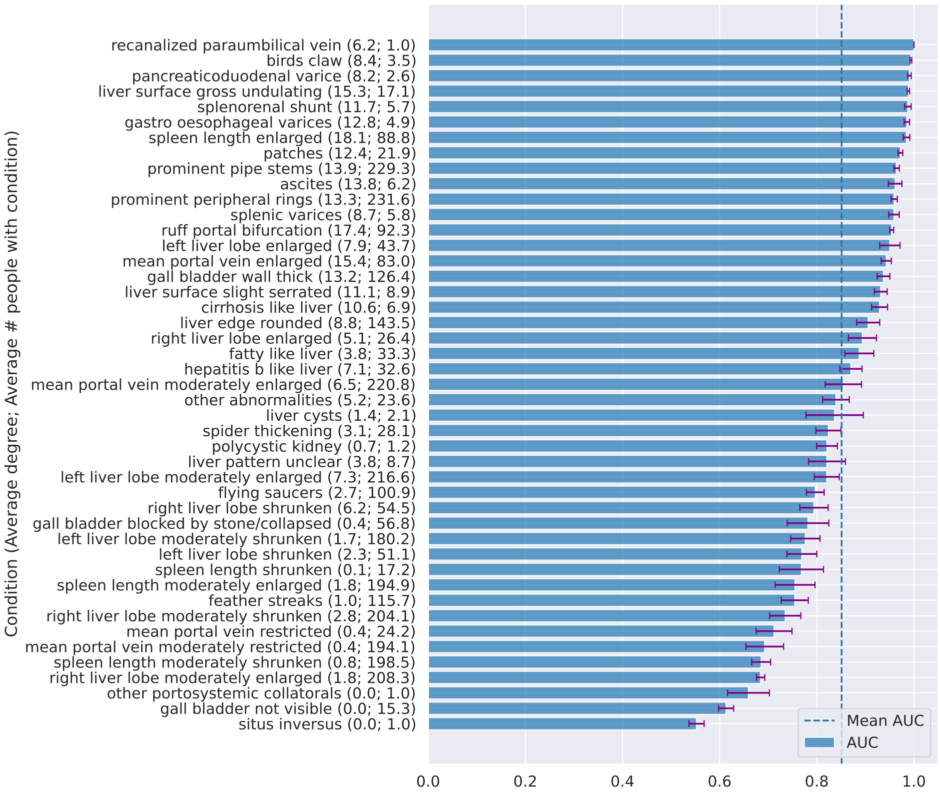


(b)

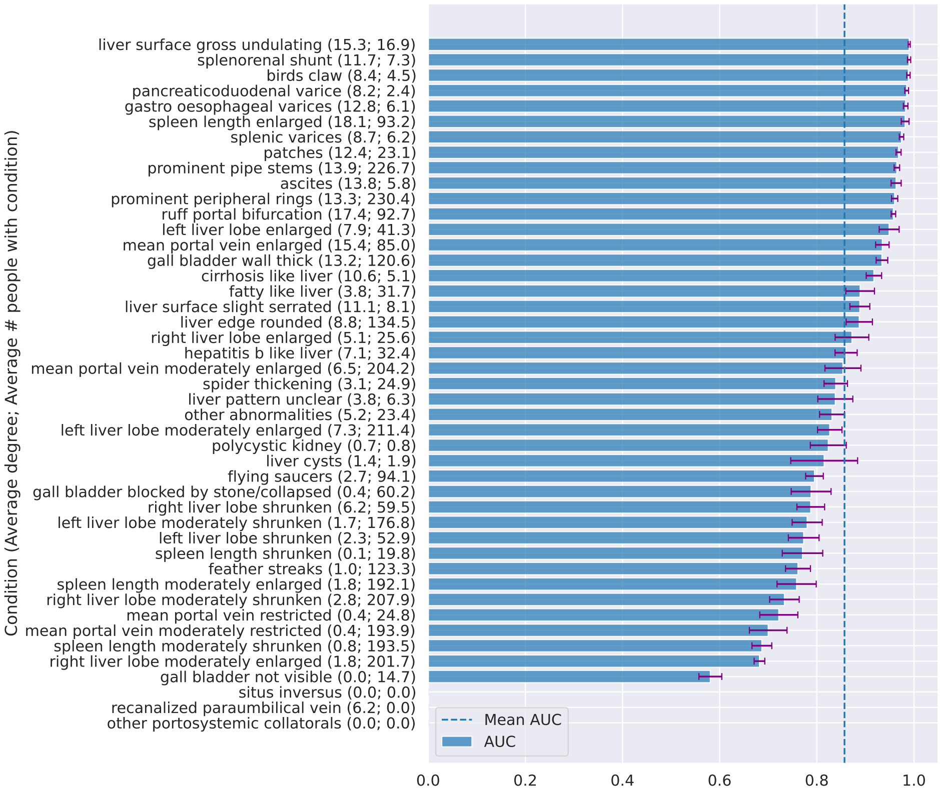


Figure S20: Performance on each condition ordered by AUC with GCN and signed distance correlation.
These result add information to Fig. 5. AUCs cannot be computed when the ground truth only consist of 1 class (when 0 people exhibited the condition) and therefore does not exist for some conditions on the test set. (a) training set (b) testing set.

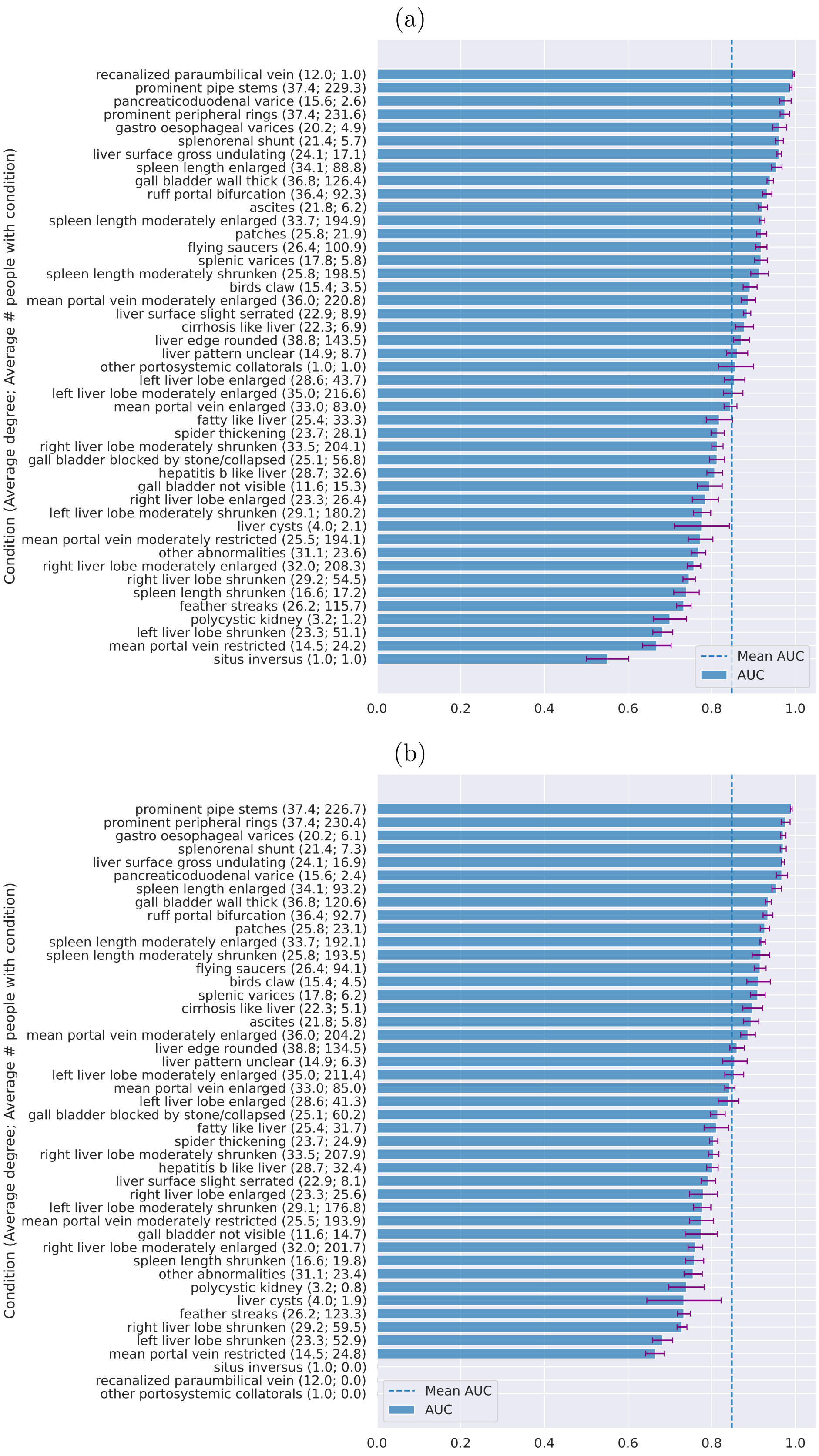


Figure S21: Performance on each condition ordered by AUC with GCN and co-occurrence.

These results add information to Fig. 5. AUCs cannot be computed when the ground truth only consist of 1 class (when 0 people exhibited the condition) and therefore does not exist for some conditions on the test set. (a) training set (b) testing set.

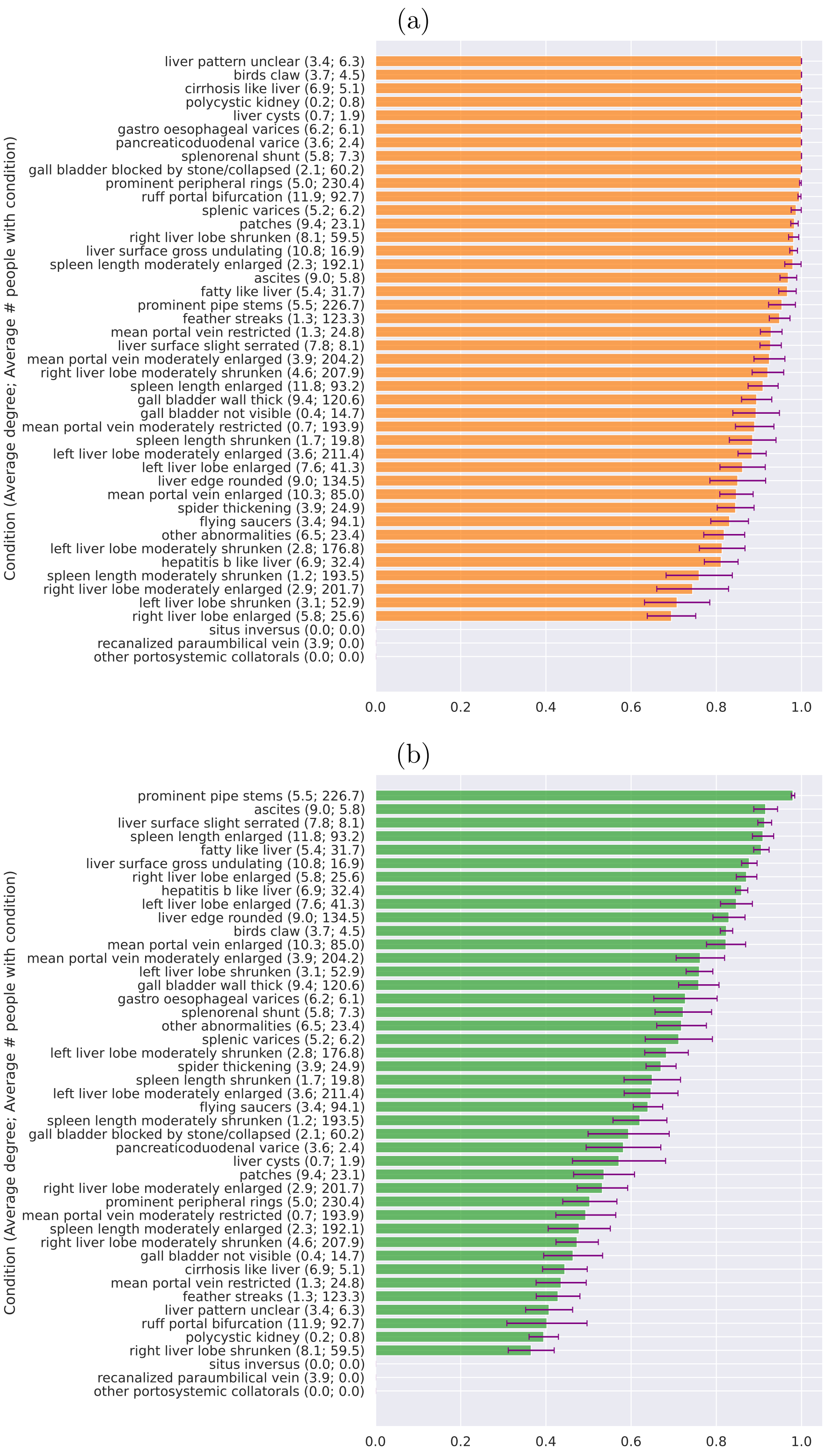


Figure S22: Performance on each condition ordered by sensitivity and specificity of GCN and graphical lasso.
The probability cut-off was chosen based on the highest sensitivity plus specificity over all conditions excluding unobserved conditions shown in Fig. S25 (a) Sensitivity (b) Specificity.

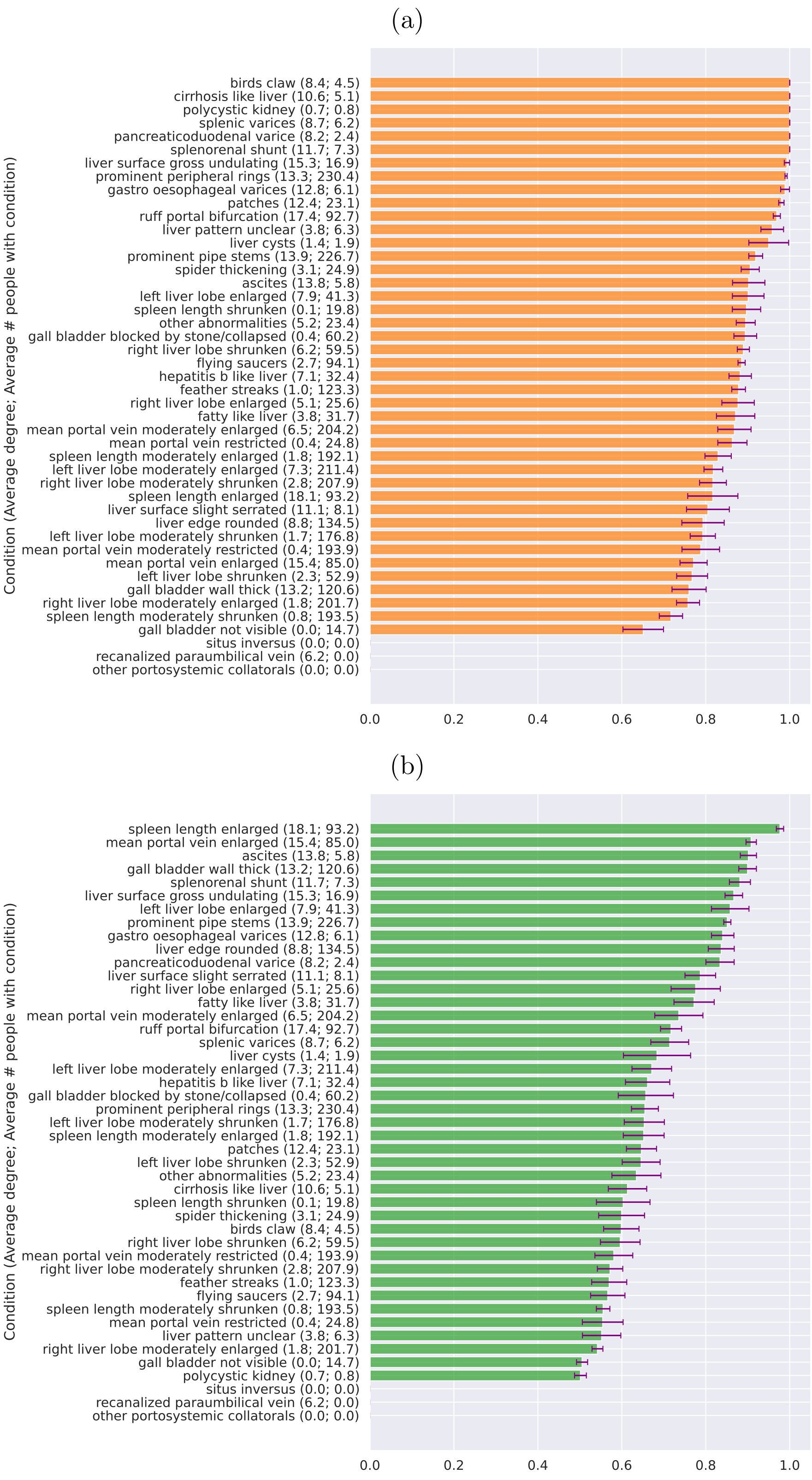


Figure S23: Performance on each condition ordered by sensitivity and specificity analysis of GCN and signed distance correlation.

The probability cut-off was chosen based on the highest sensitivity plus specificity over all conditions excluding unobserved conditions shown in Fig. S26 (a) Sensitivity (b) Specificity.

(a)

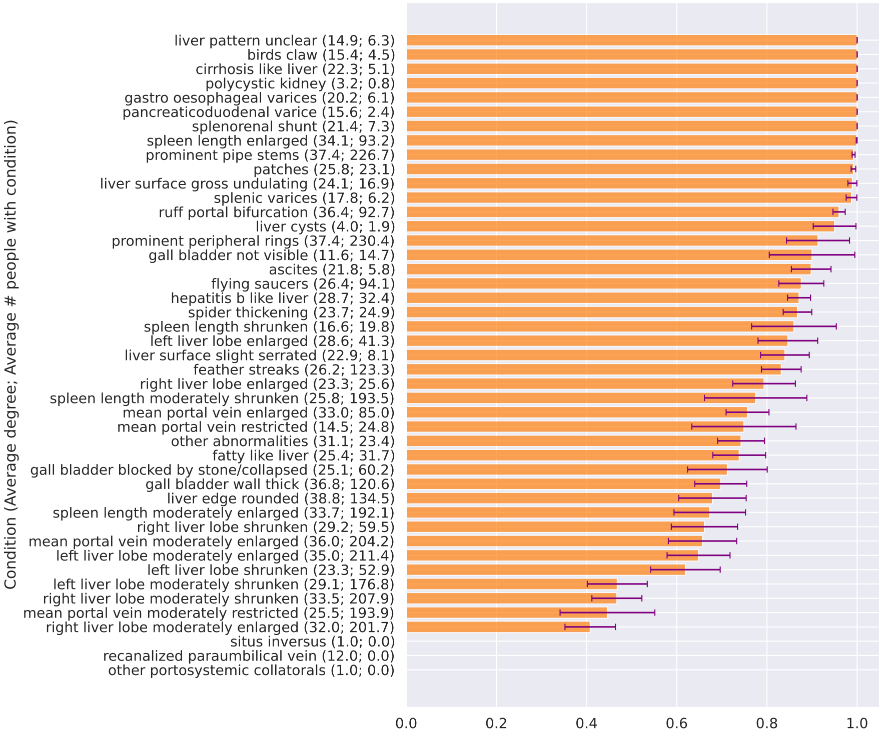


(b)

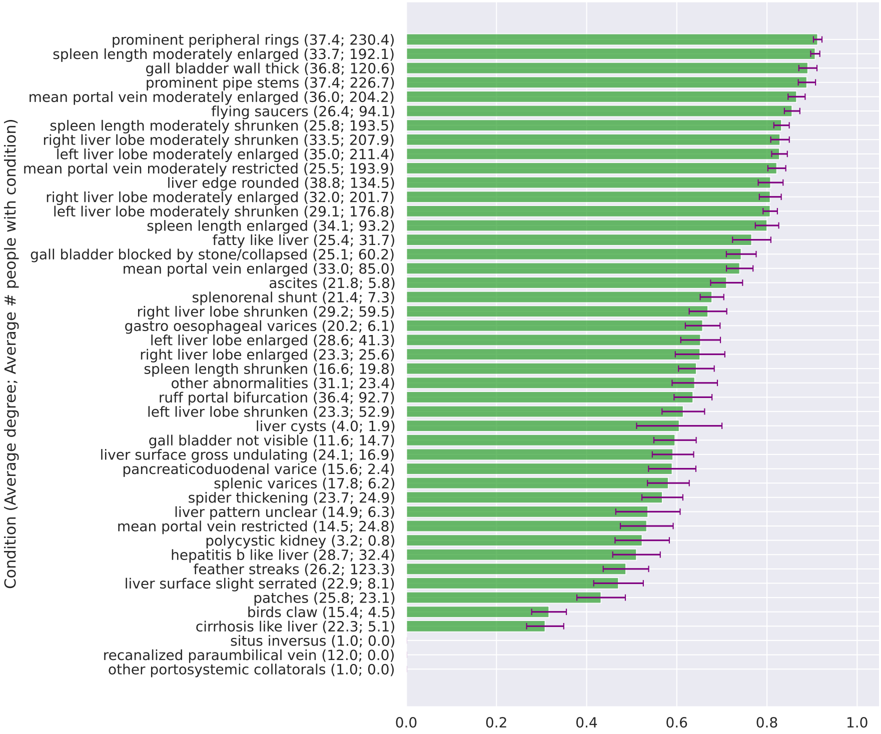


Figure S24: Performance on each condition ordered by sensitivity and specificity analysis of GCN and co-occurrence.
The probability cut-off was chosen based on the highest sensitivity plus specificity over all conditions excluding unobserved conditions shown in Fig. S27 (a) Sensitivity (b) Specificity.

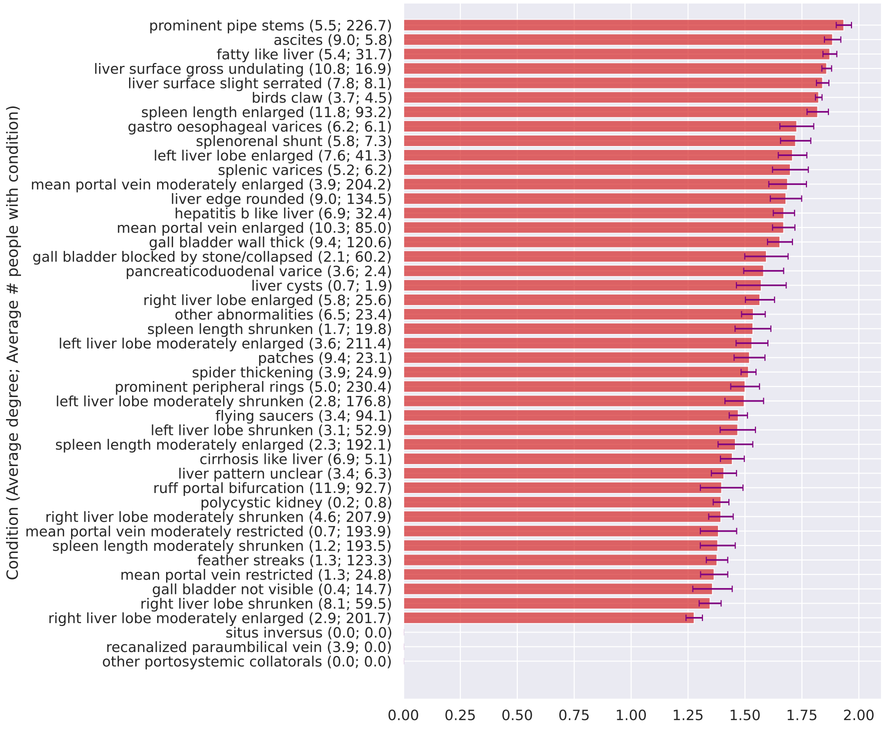


Figure S25: Performance on each condition ordered by sensitivity plus specificity of GCN and graphical lasso.
The probability cut-off was chosen based on the highest sensitivity plus specificity over all conditions excluding unobserved conditions.

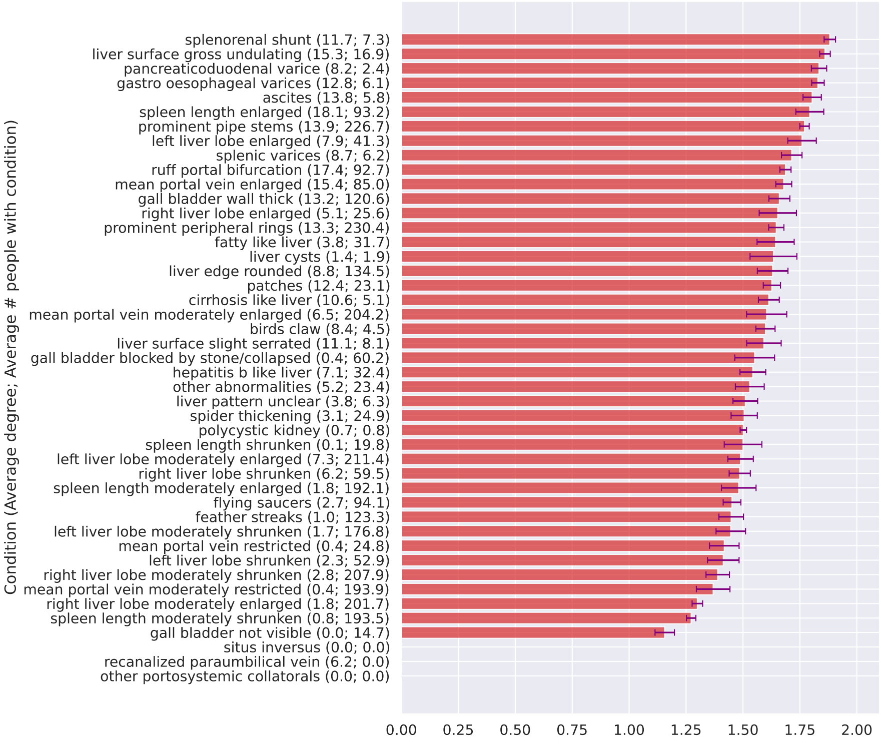


Figure S26: Performance on each condition ordered by sensitivity plus specificity of GCN and signed distance correlation.
The probability cut-off was chosen based on the highest sensitivity plus specificity over all conditions excluding unobserved conditions.

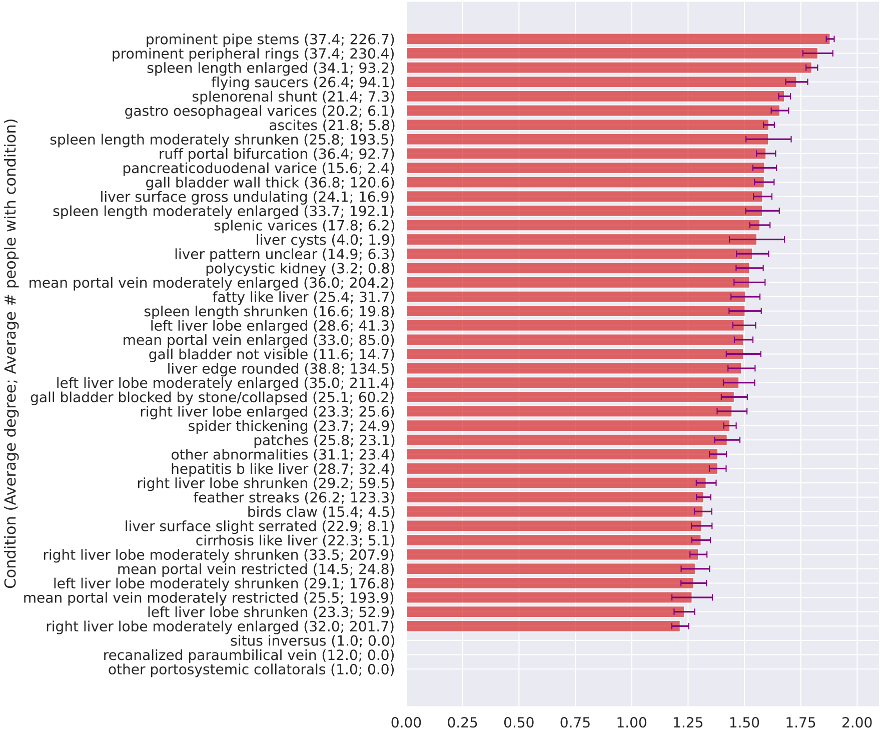


Figure S27: Performance on each condition ordered by sensitivity plus specificity of GCN and co-occurrence.
The probability cut-off was chosen based on the highest sensitivity plus specificity over all conditions excluding unobserved conditions.

### References


1 Anjorin S, Nabatte B, Mpooya S, Tinkitina B, Opio CK, Kabatereine NB, et al. The epidemiology of periportal fibrosis and relevance of current Schistosoma mansoni infection: a population-based, cross-sectional study. medRxiv. 2023:2023-09.

2 Richter J, Hatz C, Campagne G, Bergquist N, Jenkins JM, et al. Ultrasound in schistosomiasis: a practical guide to the standard use of ultrasonography for assessment of schistosomiasis-related morbidity: Second international workshop, October 22-26 1996, Niamey, Niger. World Health Organization; 2000.

3 Shervashidze N, Schweitzer P, Van Leeuwen EJ, Mehlhorn K, Borgwardt KM. Weisfeiler-lehman graph kernels. Journal of Machine Learning Research. 2011;12(9).

4 Huang NT, Villar S. A short tutorial on the weisfeiler-lehman test and its variants. In: ICASSP 2021-2021 IEEE International Conference on Acoustics, Speech and Signal Processing (ICASSP). IEEE; 2021. p. 8533-7.

5 Kriege N, Mutzel P. Subgraph matching kernels for attributed graphs. arXiv preprint arXiv:12066483. 2012.

6 Hido S, Kashima H. A linear-time graph kernel. In: 2009 Ninth IEEE International Conference on Data Mining. IEEE; 2009. p. 179-88.

7 Kipf TN, Welling M. Semi-supervised classification with graph convolutional networks. arXiv preprint arXiv:160902907. 2016.

8 Veličković P, Cucurull G, Casanova A, Romero A, Lio P, Bengio Y. Graph attention networks. arXiv preprint arXiv:171010903. 2017.

9 Hamilton W, Ying Z, Leskovec J. Inductive representation learning on large graphs. Advances in neural information processing systems. 2017;30.
